# Mapping genetic effects on splicing in ten thousand post-mortem brain samples reveals novel mediators of neurological disease risk

**DOI:** 10.1101/2025.09.25.25336663

**Authors:** Aline Réal, Kailash BP, Winston H. Dredge, Benjamin Z. Muller, Beomjin Jang, Alex Tokolyi, Hong-Hee Won, Jack Humphrey, Towfique Raj, David A. Knowles

## Abstract

Alternative splicing shapes isoform diversity and gene dosage but how genetic variation impacts splicing in brain disease is still not fully characterized. We assembled BigBrain, a multi-ancestry resource of 10,725 bulk RNA-seq profiles with matched genotypes from 4,656 individuals across 43 tissue-cohort pairs, and mapped 68,358 *cis*-sQTLs affecting 10,966 genes using mixed-model meta-analysis. Using SuSiE, we finemapped over half of these sQTLs into 95% credible sets, frequently to a single variant near splice sites. We further annotated variants predicted to alter dosage through frameshifts or nonsense-mediated decay or disrupt protein domains. Colocalization with seven neurodegenerative and psychiatric GWAS highlighted 97 loci where alternative splicing appears to mediate genetic risk. Among sQTL-eQTL pairs with colocalization probability ≥ 0.8 (posterior probability of a shared causal variant), half shared credible-set variants, showing that splicing can complement or act independently of expression. Mechanistic examples include *CAMLG* (Parkinson’s), *ZDHHC2* (Schizophrenia), and *CLU* (Alzheimer’s).

## Introduction

Alternative splicing (AS) enables one gene to produce multiple mRNA isoforms, expanding protein diversity and tuning gene expression^1^; it also adjusts gene dosage via nonsense-mediated decay (NMD)^2^. Over 95% of human genes undergo AS^3^, and the brain depends on it for glial and neuronal specialization^4,5^. Splicing defects are linked to disease, including disorders of RNA-binding proteins (RBPs) and splicing regulators^6–8^. However, how inherited variation shapes brain splicing and influences disease risk remains poorly defined, largely due to modest sample sizes to date.

Genome-wide association studies (GWAS) have identified many genetic variants linked to neurodegenerative and psychiatric disorders^9–21^, mostly in non-coding regions, complicating mechanistic interpretation. These variants are enriched in *cis*-regulatory elements (CREs) of disease-relevant cell types^22–24^, suggesting regulatory rather than coding effects. Although fine-mapping narrows candidate causal variants^25,26^, linking them to target genes remains challenging. Expression quantitative trait loci (eQTLs) map genetic variants to gene expression changes and have been used to infer mechanisms for many GWAS loci^27–31^. However, eQTLs explain only a fraction of disease heritability^32^; even in the most relevant cellular context for a given disease, such as immune cells, only ∼25% of immune-related GWAS loci can be linked to *cis*-eQTLs^33^. Recent studies highlight alternative splicing (AS) as an additional regulatory layer, with many disease variants acting through splicing rather than total expression^34–36^. Splicing QTL (sQTL) mapping identifies variants that alter splicing patterns, i.e., isoform usage, with consequences for protein abundance, structure, and function; yet sQTLs remain less studied than eQTLs, and their disease relevance is still being fully elucidated^37,38^.

Multiple studies show sQTLs contribute to disease risk independently of eQTLs^30,34,39^, despite partial overlap in regulatory effects^40^. In Alzheimer’s disease (AD), a *cis*-sQTL at *CD33* reduces exon 2 inclusion, generating a shorter isoform lacking the extracellular sialic-acid-binding domain and associating with reduced AD risk^41^, likely via diminished microglial inhibition and enhanced amyloid-β clearance. This illustrates how genetic variation can modulate splicing to alter protein function and disease susceptibility, underscoring the value of sQTL studies for uncovering mechanisms and therapeutic targets in neurodegenerative and psychiatric disorders.

Most brain sQTL studies are underpowered, typically including fewer than 1,000 individuals (e.g., GTEx v8 has ≤255 samples per brain region)^30^ limiting discovery and GWAS interpretation. A recent advance by Qi et al. mapped sQTLs in 2,865 cortex samples from 2,443 individuals^36^, underscoring the power of large-scale analyses. This study was cortex-only, European-focused, and lacked cross-tissue resolution and ancestry-aware modeling. A comprehensive brain sQTL resource that matches modern GWAS resolution and integrates ancestry, tissue diversity, fine-mapping, and functional interpretation remains unmet.

To address this, we assembled BigBrain, the largest and most diverse human brain QTL resource to date, comprising 10,725 bulk RNA-seq samples from 4,656 individuals from 11 cohorts. Beyond its scale, BigBrain is uniquely powered for discovery: it captures population diversity, harmonizes brain phenotypes across studies, and integrates matched genotypes, enabling analyses that individual datasets cannot support. This scale enables the detection of smaller-effect and lower-frequency variants, while supporting high-resolution fine-mapping and mechanistic dissection of splicing regulation. Using a linear mixed model meta-analysis framework^42^, which accounts for sample structure and relatedness, we mapped *cis*-sQTLs, then performed statistical fine-mapping and GWAS colocalization to identify splicing events likely mediating genetic risk. Our analyses uncovered numerous fine-mapped sQTLs and sQTLs that colocalize with disease loci, implicating unproductive splicing and domain disruption. These results reveal how disease-associated variants perturb isoform usage and protein architecture, linking genetic risk to molecular function.

Importantly, BigBrain fills a critical gap by providing a large-scale, brain-specific, and mechanistically annotated splicing QTL resource at higher resolution. BigBrain is built as a community resource that enables researchers to (i) prioritize causal variants and transcripts at GWAS loci, (ii) generate mechanistic hypotheses for experimental validation, and (iii) benchmark splicing-aware analytical and machine-learning methods. Altogether, our work shows that alternative splicing is a widespread, underappreciated mediator of genetic risk in the brain, and BigBrain provides a powerful resource for elucidating the genetic and molecular basis of neurological disorders.

## Results

### RNA-seq harmonization for splicing phenotypes

We assembled ‘BigBrain’ by integrating paired RNA-seq and genotype data from 11 cohorts (43 tissue-cohort pairs), totaling 10,725 samples from 4,656 donors spanning different tissues from five major brain regions (**Fig. 1a, Supplementary Figure 1a**). The cohort includes individuals from multiple ancestries: 83.3% European, 12.1% African, 3.5% Hispanic and < 1% East and South Asian (**Fig. 1b, Supplementary Figure 1b, Table 1**).

**Figure 1.**
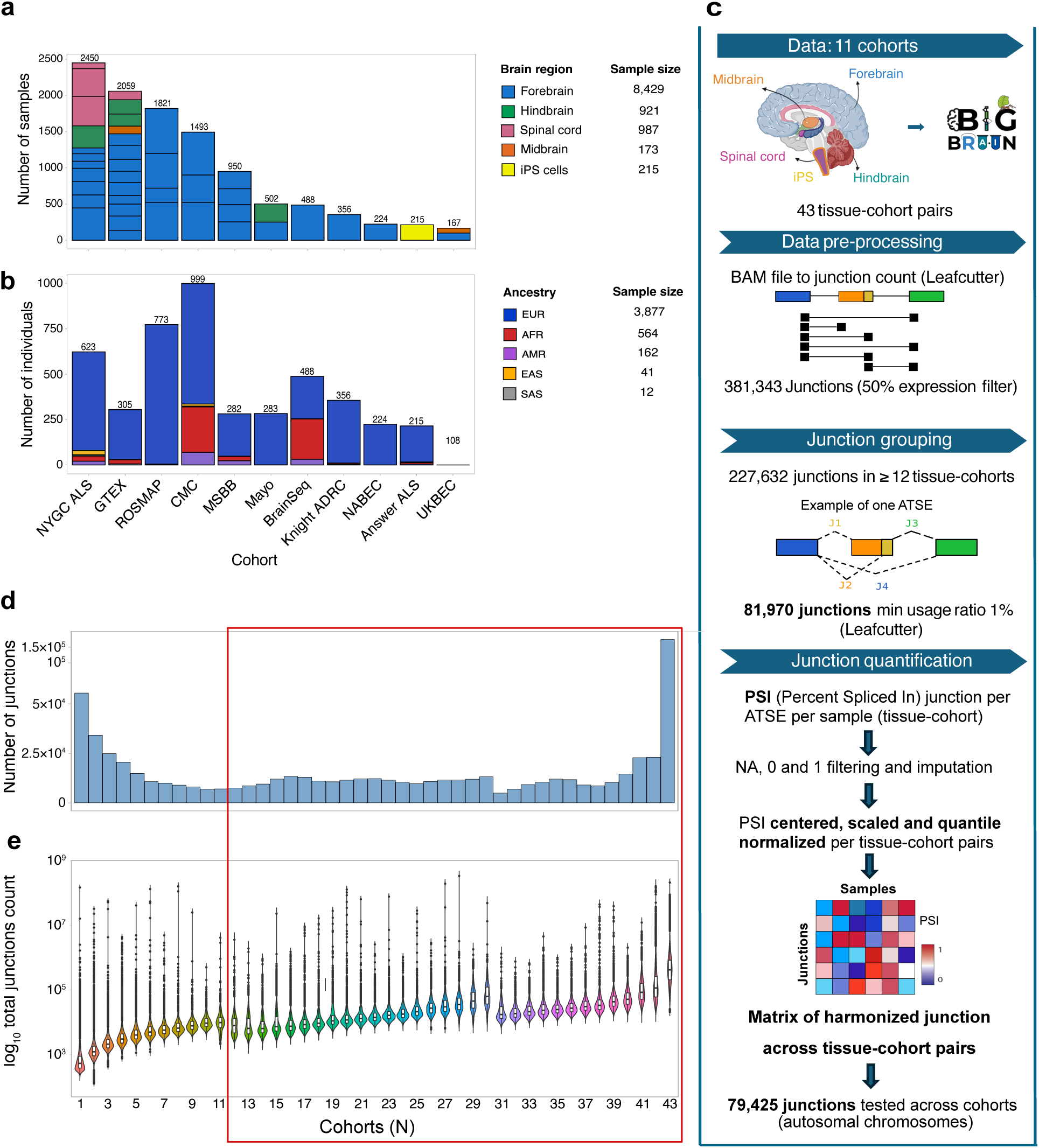
Cohort overview and junction harmonization pipeline in the BigBrain dataset. **(a)** Bar plot showing the distribution of RNA-seq samples per tissue-cohort pair across brain regions and iPSC-derived neurons. Each bar is colored by tissue type. **(b)** Stacked bar plot showing the inferred ancestry breakdown per cohort, with contributions from European (EUR), African (AFR), Admixed American (AMR), East Asian (EAS), and South Asian (SAS) populations. **(c)** Schematic diagram of the splicing quantification pipeline. Junctions were extracted using Leafcutter, grouped into alternative transcript splicing events (ATSEs), and harmonized using normalized PSI values. **(d)** Bar plot showing the number of retained junctions per tissue-cohort pair after expression filtering. The x-axis represents tissue-cohort pairs. **(e)** Violin plot showing the distribution of log_10_-transformed total junction counts per sample across all 43 tissue-cohort combinations. The x-axis denotes tissue-cohort pairs; the y-axis shows log_10_ total junction count per sample.

Harmonizing splicing junctions across tissues, populations, and cohorts is critical in large-scale multi-cohort studies. In BigBrain we used LeafCutter^43^, which identifies splicing junctions by leveraging spliced reads spanning introns to quantify intron usage across samples and build a junction-level harmonization pipeline (**Fig. 1c**). To avoid including rare or tissue-specific junctions confined to a few cohorts, we required cross-cohort support: a junction had to be present in ≥ 50% of samples within a cohort and meet this threshold in ≥ 12 of the 43 pairs, yielding 227,632 junctions (**Fig. 1d**). These junctions accounted for the majority of the variance across tissue-cohort pairs in intron usage and are more highly expressed across cohorts (**Fig. 1e**). Differences in RNA-seq library preparation protocols (e.g., strandedness and read length) can introduce variability in splice junction detection. To address this, we applied unified junction extraction, clustering and normalization, enabling consistent cross-dataset comparability and correction of technical variation identified through principal component analysis (PCA) (**Materials and Methods**). To our knowledge, BigBrain is the largest junction-level harmonized brain RNA-seq resource, enabling consistent splicing event definitions and cross-sample comparability at unprecedented scale.

We grouped junctions with LeafCutter jointly across all tissue-cohort pairs to ensure consistent event definitions (**Fig. 1c**). We defined Alternative Transcript Structure Events (ATSEs; also known as clusters) as groups of splice junctions sharing splice sites, derived from split-read evidence, that capture alternative splicing including alternative first/last exons. ATSEs offer a unified framework to analyze transcript structure diversity (ATSE example, middle panel **Fig. 1c**). Using a 1% minimum usage, we detected 26,878 multi-junction ATSEs (median 3, max 34) spanning 81,970 junctions (79,425 within autosomes) (**Supplementary Figure 1c-d**). For each junction, we computed Percent Spliced In (PSI) as the fraction of reads supporting that junction within its ATSE; the resulting 79,425 × samples PSI matrix was quantile-normalized per tissue-cohort pair for downstream sQTL discovery (**Fig. 1c, bottom panel**).

To assess the overall variance structure in PSI values, we performed PCA (diagnostic only) and quantified the proportion of variance explained by the first 50 principal components (PCs). The top six PCs explained > 50% of variance, reflecting strong underlying biological and technical influences (**Supplementary Figure 1e**). PC2 (adj. R² = 0.899), PC5 (0.893), and PC1 (0.65) captured major cohort and brain-region effects (**Supplementary Figure 1f-h)**. Technical factors like library preparation, strand specificity, age, loaded mainly on lower PCs (**Supplementary Figure 1i-j**).

### Multi-ancestry mixed model for *cis*-sQTL meta-analysis

After stringent junction filtering and PSI harmonization, we mapped multi-ancestry *cis*-sQTLs across all 43 tissue-cohort pairs by meta-analyzing SNPs within ±1 Mb of each junction body (start to end). We used mmQTL^42^, a linear mixed-model meta-analysis that accounts for population structure (∼16% non-European ancestry), relatedness, and repeated donors (e.g., multiple brain regions per individual in GTEx and NYGC ALS Consortium (*Humphrey et al., in preparation*)). Our model included age, sex, and 15 PEER factors^44^ to control technical confounding. We chose PEER over PCs because PEER is designed to capture hidden confounders (batch, RNA quality, cross-cohort effects) while better preserving local genetic signal; in diagnostics, PEER factors correlated with the leading PSI PCs (data not shown), indicating they absorb the same major unwanted variation.

We mapped *cis*-eQTLs and *cis*-sQTLs in BigBrain to quantify genetic regulation of expression and splicing. In the multi-ancestry meta-analysis (n = 4,656), we detected 22,663 eGenes, genes with a significant *cis*-eQTL at FDR < 5%, and 68,357 lead sQTLs (top association per junction at FDR < 5%) spanning 24,064 ATSEs, 10,966 sGenes, and 86% of tested junctions. Restricting to individuals from European genetic ancestry (n = 3,877) yielded 22,431 eGene and 68,062 lead sQTLs (FDR < 5%) across 24,130 ATSEs and 10,981 sGenes; 85.6% of tested junctions had a significant lead association (**Fig. 2a**). We observed that sQTL discovery scales with sample size using the average squared Z-scores (Z²) (**Supplementary Figure 2a**). Although mmQTL adjusts for population structure, it implicitly assumes shared effect sizes across ancestries. In reality, splicing effects can differ by ancestry owing to allele-frequency, linkage disequilibrium (LD), and regulatory differences, which likely explains why pooling all ancestries did not increase sQTL yield over the European-only analysis.

**Figure 2.**
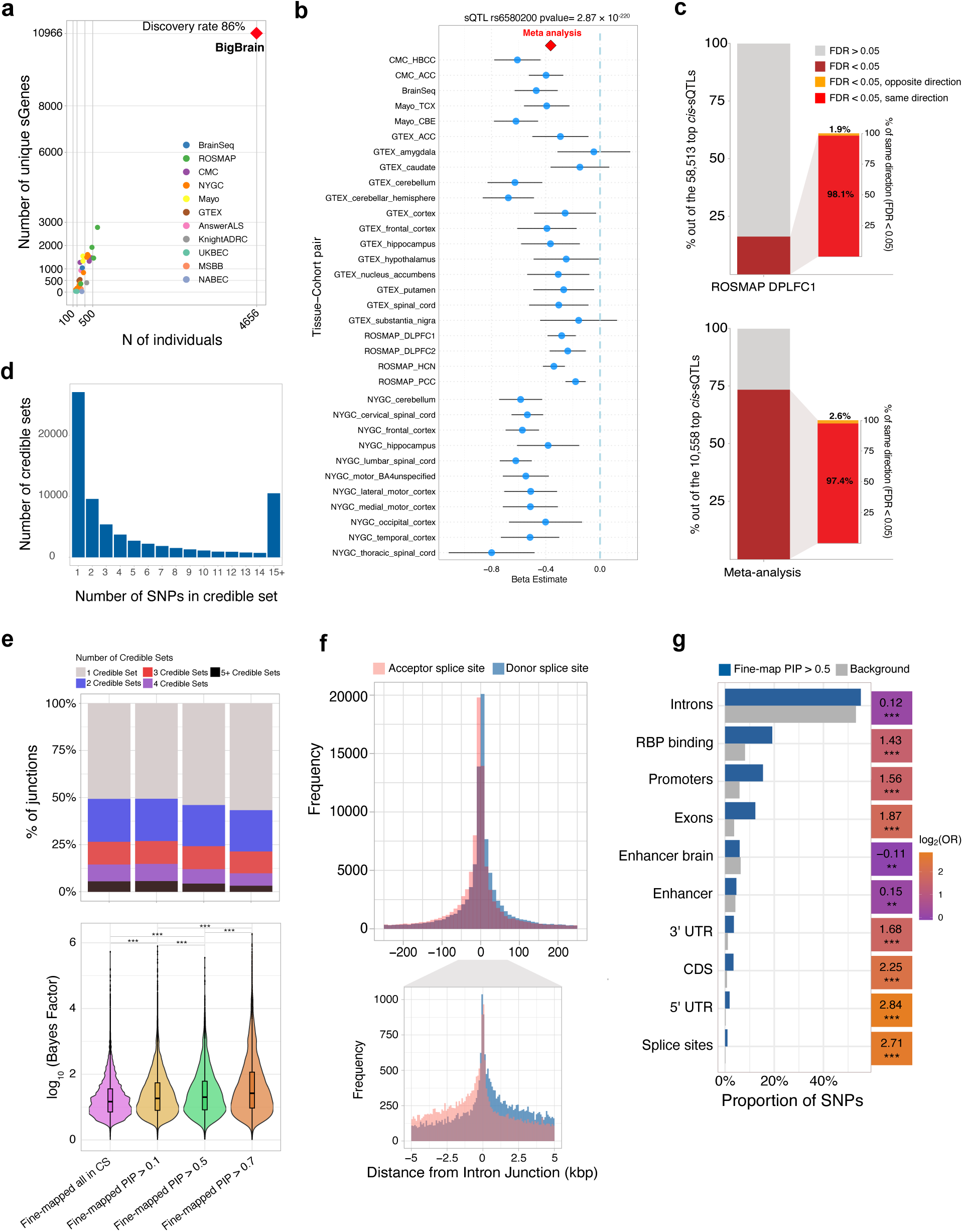
sQTL discovery, replication, and fine-mapping in the BigBrain dataset. **(a)** Number of unique sGenes (gene affected by a significant sQTL) identified per cohort as a function of sample size in all multi-acestries part of the BigBrain dataset. BigBrain meta-analysis shows the highest discovery rate compared to single-cohort analysis. **(b)** Forest plot showing beta estimates and 95% confidence intervals for the splicing QTL rs6580200 across tissue-cohort pairs. The effect size for each cohort is represented by a blue dot with a horizontal line indicating the confidence interval. The red diamond denotes the overall meta-analysis estimate (*P* = 2.87 × 10^-220^), demonstrating consistent effects across multiple brain regions. **(c)** Replication of top *cis*-sQTLs mapped in a held-out cohort (ROSMAP DLPFC, N=523) into the meta-analysis without the held-out cohort and vice-versa. Bars indicate sQTLs with consistent direction and significance. **(d)** Distribution of the number of SNPs per credible set for fine-mapped sQTLs. **(e)** Top: Proportion of junctions with 1, 2, or ≥ 3 credible sets across different PIP thresholds. Bottom: Distribution of fine-mapping evidence strength across different fine-mapping PIP filters (all in 95% CS, and those with PIP > 0.1 / 0.5 / 0.7). The y-axis shows log_10_(Bayes factor) (per-variant; SuSiE LBF). Violins depict the density; the box marks median and IQR (whiskers = 1.5 × IQR). Higher values indicate stronger support for causality. Horizontal brackets show pairwise tests; ***** = BH-FDR < 0.001 (two-sided Wilcoxon rank-sum). **(f)** Distribution of fine-mapped SNPs relative to donor and acceptor splice sites, showing enrichment near splice junctions. **(g)** Enrichment of fine-mapped SNPs (PIP > 0.5) across genomic annotations compared to background SNPs, with log odds ratios.

Our high *cis*-sQTL discovery underscores the power of large sample size and cross-cohort meta-analysis: we detect an 86% discovery rate, exceeding GTEx v8 (∼60-75% across tissues/approaches)^2,30^, likely reflecting both scale and the complexity of brain splicing. To enable downstream comparisons with eQTLs, RNA editing QTLs (edQTLs)^45^, and GWAS, we tested SNPs within ±1 Mb of each junction; although some sQTL studies use ±100 kb^2,30^, the wider window aligns with other molecular QTL/GWAS analyses and still captures splice-site-proximal enrichment (**Supplementary Figure 2b**). Because meta-analysis boosts power but requires heterogeneity assessment, we compared *cis*-sQTL effect sizes (β) across BigBrain for shared SNP-junction pairs. We observed concordance within similar cohorts/tissues (e.g., within ROSMAP and CommonMind Consortium (CMC)) and across spinal cord tissues (**Supplementary Figure 3c**). Despite variability in SNP coverage, shared pairs showed strong correlations, notably NYGC vs Mayo cerebellum (Pearson R = 0.97, p < 2×10⁻¹⁶) and NYGC-ALS vs GTEx spinal cord (R = 0.96, p < 2×10⁻¹⁶) (**Supplementary Figure 2c-d**). As an illustrative example, rs6580200 is among our strongest *cis*-sQTL signals (*P* = 2.87×10⁻²²⁰), regulating *CXXC5* (CXXC-type zinc finger protein 5) splicing across multiple brain regions (**Fig. 2b**). Prior work suggests this variant disrupts an *HNRNPK* (heterogeneous nuclear ribonucleoprotein K) binding site, with region-specific effects potentially linked to high *HNRNPK* expression in the cerebellar hemisphere^46^; the locus also emerges as a top sQTL candidate in SCZ.

To assess *cis*-sQTL replication, we applied Storey’s π₁ in an independent held-out cohort (ROSMAP-DLPFC1, N = 522). Of the top 58,513 *cis*-sQTLs from a meta-analysis excluding ROSMAP-DLPFC1, 16.3% replicated at FDR < 5% in the held-out, consistent with prior sQTL studies, and 98% showed concordant effect directions. Conversely, testing significant ROSMAP-DLPFC1 sQTLs against the meta-analysis (excluding ROSMAP-DLPFC1) yielded 73% replication (FDR < 5%) with 97.4% directional concordance, an asymmetry driven by the smaller holdout (**Fig. 2c**).

### Bayesian fine-mapping of *cis*-sQTLs with SuSiE identifies putative causal variants

To identify putative causal variants underlying splicing regulation, we next applied Bayesian fine-mapping with SuSiE^47,48^. From 68,063 European *cis*-sQTLs, SuSiE fine-mapped 52.7% (35,879 junctions) to at least one credible causal variant; 657,309 SNP-junction pairs were assigned to 95% credible sets (**Supplementary Figure 2e** shows genome-wide PIPs as a Manhattan-like plot). Importantly, 50.7% of credible sets were single-variant (**Fig. 2d**), whereas large sets (> 15 variants) had low support (median PIP ≈ 0.01), indicating lower resolution and weaker signal confidence.

At the junction level, 50.7% of fine-mapped junctions mapped to a single credible set and 49.3% to ≥ 2; this proportion held at PIP > 0.1 and shifted toward single signals with stricter thresholds (53.9% vs. 46.1% at PIP > 0.5 and 56.7% vs. 43.3% at PIP > 0.7), with some junctions showing up to five independent sets (**Fig. 2e Top; Supplementary Figure 2f**). As PIP thresholds increase, lower-confidence associations are filtered, enriching for single-signal loci. To assess signal strength, we compared log_10_ Bayes factors (logBF) across PIP bins (**Fig. 2e Bottom**). logBF increased with PIP, consistent with stronger causal evidence at higher PIPs. All pairwise contrasts were significant (e.g., PIP 0.5-0.7 vs 0.1-0.5: *P* = 1.9×10⁻⁶; all other comparisons: *P* < 2×10⁻¹⁶), supporting the complementary use of PIP and logBF.

Fine-mapped *cis*-sQTLs were strongly positionally enriched around splice junctions, peaking at donor and acceptor sites (**Fig. 2f**); most fell within ±100 bp. Functionally, they were significantly enriched in exons, splice sites, UTRs, promoters, and RNA-binding protein (RBP) binding sites (**Fig. 2g**), consistent with disruption not only of canonical splice elements, but also RBP-mediated regulation, and transcriptional-splicing coupling^49,50^. Conversely, they were depleted in brain-specific and generic enhancers (**Fig. 2g**), reinforcing the specificity of fine-mapped *cis*-sQTLs for splicing-related mechanisms.

### Impact of sQTL-affected junctions on protein domains

To characterize the genomic context of splice junctions, we mapped all tested junctions to annotated splice sites using leafidex, classifying them as donor, acceptor, or both-site supported (**Supplementary Figure 4a**). Significant *cis*-sQTL-regulated junctions (sJunctions) were then overlapped with exons, coding sequence (CDS), and UniProt protein domains within a ±10 bp window. Overall, 88% of sJunctions overlapped exons, 71.5% CDS, and 26% protein domains (**Fig. 3a, left pane**l).

**Figure 3.**
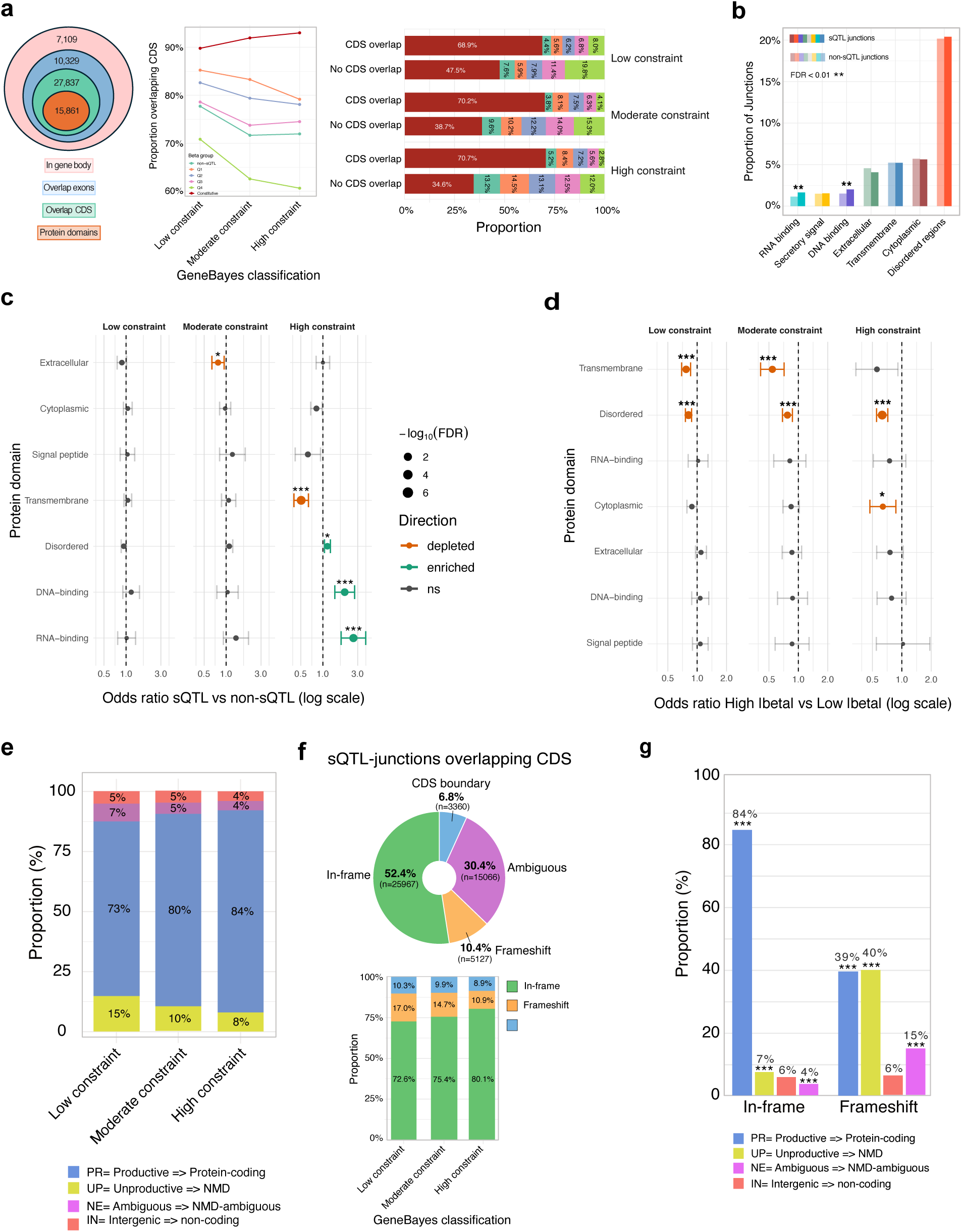
Functional impact of sQTL-affected junctions on protein domains. **(a)** Left: Venn diagram showing the number of significant sQTLs affected junctions (sJunctions) overlapping gene body, exons, coding sequence (CDS), and protein domains, annotated using the *leafidex* package and UniProt annotations. Middle: Proportion of sJunctions overlapping CDS across GeneBayes constraint tiers, stratified by effect-size bin (β quartiles Q1–Q4), non-sQTL, and constitutive junctions. Right: For each GeneBayes tier, stacked bars give the composition of junction categories among CDS-overlap (top) vs No CDS overlap (bottom): non-sQTL, sQTL effect-size bins Q1–Q4, and constitutive (cluster-dominant) junctions. Values are within-bar percentages. **(b)** Proportion of junctions overlapping different protein domain categories in sQTLs (darker colors) vs. non-sQTLs (lighter colors). **(c-d)** χ² screen for domain enrichment across gene-constraint tiers. Within each GeneBayes tier (Low/Moderate/High), we built 2×2 tables for each protein-domain class: **c**: domain × (sQTL vs non-sQTL) and **d**: domain × (high-|β| vs low; non-sQTLs set to β = 0; high = top decile within run). Points show odds ratios (OR; log scale) comparing the domain to all other junctions in the same tier; bars = 95% CI. Point size encodes −log₁₀(FDR) (capped at 10); color indicates direction (green = enrichment, orange = depletion, grey = ns). Asterisks mark FDR < 0.05. P-values come from Pearson’s χ² (Fisher’s exact when expected counts < 5). The vertical dashed line marks OR = 1. **(e)** Stacked bars show the proportion of sJunctions classified as productive (PR; protein-coding), unproductive (UP; NMD), ambiguous (NE; NMD-ambiguous), and intergenic (IN; non-coding) in Low, Moderate, and High GeneBayes constraint groups. Percent labels are within-tier shares; colors match the legend below. **(f)** Pie-chart for coding consequences of sJunctions overlapping CDS regions. Junctions were classified as in-frame (50.9%), frameshift (20.2%), or ambiguous (25.6%) and CDS boundary (3.3%) based on CDS overlap. **(g)** Proportion of sJunctions by frameshift status and predicted splicing productivity class. Frameshift-inducing junctions were significantly enriched in unproductive (UP) and ambiguous (NE) categories compared to in-frame junctions (Fisher’s exact test; \*\*\**P* < 0.001).

Restricting to CDS-overlapping sJunctions, we stratified by absolute sQTL effect (|β| quartiles Q1–Q4) and GeneBayes constraint (low/moderate/high by quantile bins)^51^. Across β bins, odds ratios were < 1 in high-constraint genes versus low, with the strongest depletion for Q4 (largest effects sQTLs) and constitutive junctions increased with gene constraint (**Fig. 3a, right**). This pattern suggests that selection removes large splicing effects in dosage-sensitive genes, while moderate effects persist, balancing function and regulatory flexibility. Within each GeneBayes tier, the Q4 fraction was significantly reduced among CDS-overlapping junctions, most prominently in high-constraint genes, whereas constitutive junctions increased with constraint (**Fig. 3a, right**). In line with Glassberg et al.^52^, who reported modest constraint on expression variation, our results indicate that selection dampens, but does not eliminate, large-effect splicing QTLs, allowing constrained genes to retain limited regulatory flexibility.

Comparing sJunctions vs. non-sQTL junctions across UniProt domains revealed that disordered regions were most frequently excised, while RNA- and DNA-binding domains were significantly enriched among sQTLs (**Fig. 3b**), implying greater regulatory susceptibility of these domain types.

To test the joint effects of domain context and gene constraint, unadjusted Pearson χ² tests within each GeneBayes tier (Low/Moderate/High) showed significant depletion of sQTLs in transmembrane (High) and extracellular (Moderate) regions, and enrichment in RNA/DNA-binding and disordered domains (High) (FDR < 0.05; **Fig. 3c)**. For effect size (high |β| vs. low; non-sQTL = β 0), large effects were depleted in transmembrane, disordered, and cytoplasmic regions (FDR < 0.05; **Fig. 3d**). Logistic regressions including domain class, constraint, and their interaction recapitulated these χ² trends (**Supplementary Fig. 4b-c**): transmembrane domains showed negative associations with sQTL presence, whereas RNA/DNA-binding and disordered regions were positive, strongest in high-constraint genes. Effect-size coefficients were broadly negative under moderate/high constraint, confirming depletion of large effects in membrane-associated and disordered regions. Overall, domain localization predicts where sQTLs occur, enriched in nucleic-acid-binding and disordered regions, especially in constrained genes, while constraint modulates effect magnitude, with purifying selection limiting strong splicing perturbations in dosage-sensitive contexts.

Across GeneBayes tiers, higher constraint shifted sQTL junctions toward protein-coding (PR) outcomes, with declines in unproductive (UP), ambiguous (NE), and intergenic (IN) classes (**Fig. 3e**). Stratified by sQTL status, sJunctions were enriched for non-productive/NMD-ambiguous and depleted for intergenic events, while PR proportions were similar to non-sQTLs (Fisher/χ², FDR-controlled; **Supplementary Fig. 4d**). Among CDS-overlapping sJunctions, outcomes were in-frame 52.4%, frameshift 10.4%, ambiguous 30.4%, in-frame increased with constraint, consistent with selection (**Fig. 3f; Supplementary Fig. 4e**). Linking splicing frame to translation (**Fig. 3g**), in-frame junctions were predominantly productive (PR 84%), whereas frameshifts were mostly unproductive or NMD-prone (**Supplementary Fig. 4f**), reflecting open reading frame disruption and reduced coding potential. Overall, coding-sequence sQTLs tend to preserve reading frame in constrained genes, while frameshift-inducing variants are rare and typically yield non-productive transcripts, consistent with purifying selection on translational integrity.

### *cis*-sQTL colocalization reveals splicing mechanisms underlying neurological traits

We quantified the proportion of SNP-based heritability mediated by splicing (h²_med_/h²) across several complex traits using MESC (Mediated Expression Score Regression^32^). Parkinson’s disease (PD)^19^ and bipolar disorder (BPD)^21,53^ showed the highest mediated heritability, suggesting a strong role for splicing regulation (**Fig. 4a**, **Table 3**). Notably, for PD, sQTLs explained more heritability than eQTLs, and transcript expression QTLs captured even more (**Supplementary Figure 5a**), highlighting the importance of post-transcriptional regulation in mediating genetic risk^54^.

**Figure 4.**
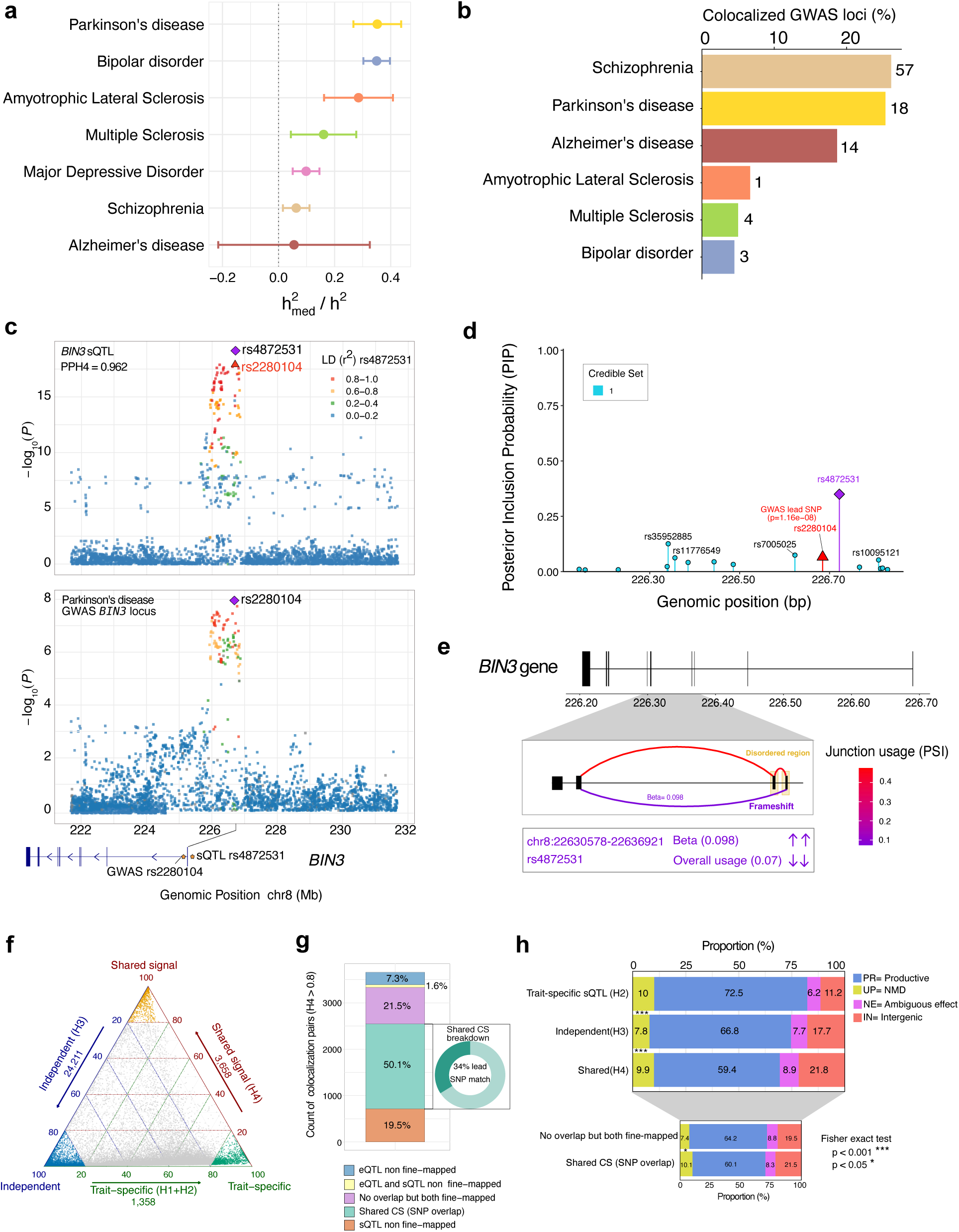
Cross-trait colocalization of sQTLs with neuropsychiatric GWAS loci and functional characterization. **(a)** Ratio of mediated heritability (h²_med) to total heritability (h²) for sQTLs across seven brain-related traits, estimated using mediated expression score regression (MESC). Error bars represent 95% confidence intervals. **(b)** Percentage of genome-wide significant GWAS loci with evidence of colocalization (PP.H4 ≥ 0.8) with sQTLs for each disorder. **(c)** Colocalization at the *BIN3* locus. Top: sQTL association (–log₁₀P) for junction chr8:226,343,632–226,366,084 in the *BIN3* gene; Bottom: GWAS association for Parkinson’s disease at the same locus. SNPs are colored by linkage disequilibrium (LD) with the lead SNP rs4872531. **(d)** Posterior inclusion probability (PIP) from SuSiE fine-mapping of sQTL for *BIN3* locus. SNPs in the 95% credible set are highlighted in cyan. The Parkinson’s GWAS is highlighted as red and part of the same credible set. **(e)** Schematic representation of the *BIN3* junction with PSI (color, upper) and sQTL beta value (box, lower). Frameshift and disordered region annotations are shown. **(f)** Ternary plot showing colocalization classification between sQTL and eQTL based on COLOC posterior probabilities. Points are colored by colocalization group: shared signal (PP.H4 ≥ 0.8), independent (PP.H3 ≥ 0.8), and trait-specific (PP.H1+PP.H2 ≥ 0.8). **(g)** Classification of colocalized sQTL-eQTL pairs (PP.H4 ≥ 0.8) based on fine-mapping results. Bars show the proportion of loci where the credible set (PIP ≥ 0.1) is shared between traits, partially mapped (credible set in only one trait), or not fine-mapped in either sQTL or eQTL. **(h)** Top: Proportion of transcript productivity classes, productive (PR), unproductive (UP), ambiguous functional effect (NE), and intergenic (IN), among sQTLs stratified by colocalization status (trait-specific, independent, shared). Bottom: Productivity class distribution among shared sQTL-eQTLs, further grouped by credible set (CS) overlap status (overlapping vs. non-overlapping). Statistical significance assessed using Fisher’s exact test (*P* < 0.05; \*\**P* < 0.001).

We colocalized significant sQTLs with these seven neurological traits using summary statistics from 14 GWAS^9–21,53^, defining high-confidence events as PP.H4 ≥ 0.8 (plus additional filters, **Methods**). SCZ had the most colocalized loci, followed by PD and AD; by proportion of loci tested, SCZ and PD were highest (**Fig. 4b**). In total, 225 junction-trait pairs colocalized, of which 178 were the top signal per junction, corresponding to 112 unique genes (**Fig. 4b**, **Table 4**). As an example, at *BIN3*, a *cis*-sQTL rs4872531 (*P* = 8.97×10⁻²⁰) located upstream of the *BIN3* gene body colocalizes with PD GWAS (PP.H4 = 0.962), suggesting a common genetic mechanism linking splicing regulation and disease risk (**Fig. 4c**). Fine-mapping identifies a single 95% credible set (17 variants) with rs4872531 top (PIP = 0.35); inclusion of the PD GWAS lead rs2280104 supports a shared causal signal (**Fig. 4d**). The sQTL promotes exon-4 skipping (predicted frameshift) and, though the least-used junction in the ATSE, shows the largest effect (**Fig. 4e**). While *BIN3* has been implicated via expression, our data indicate splicing may also contribute at this locus; rs4872531, is in strong LD with rs2280104 (r² > 0.8), nominating *BIN3* as a splicing-mediated effector in PD.

### Colocalization and fine-mapping overlap between sQTLs and eQTLs

To test whether variant effects on AS are shared with expression or distinct, we colocalized sQTLs and eQTLs using COLOC^55^. Loci were classified as shared (PP.H4 > 0.8, common causal variant), independent (PP.H3 > 0.8, distinct regulation), or phenotype-specific (PP.H1 or PP.H2 > 0.8) (**Fig. 4f, Supplementary Figure 5b**). The ternary plot (**Fig. 4f**), shows many loci near the shared vertex, indicating coordinated regulation; others fall near the independence corner or remain ambiguous. Genomic context matched these categories: sQTL-only (H2) loci were predominantly intronic, whereas shared (H4) loci were more evenly distributed across introns, exons, and promoters, suggesting partly distinct regulatory architectures (**Supplementary Figure 5c**).

COLOC assumes one shared causal variant per locus, which can miss multi-signal architectures. It can miss cases where multiple causal variants influence each trait independently, where only a subset of signals overlap, or where the shared regulation is not driven by the most strongly associated variants. We therefore compared SuSiE 95% credible sets (CS) for sQTLs and eQTLs. Among 3,658 H4 loci (PP.H4 ≥ 0.8), 50.1% shared ≥ 1 fine-mapped SNP (**Fig. 4g**), while only 34% shared the same lead SNP, underscoring the added value of CS overlap. A further 21.5% were fine-mapped for both traits but had no CS overlap, indicating independent variants. Where CS were shared, sQTL CS were significantly smaller than at non-overlapping loci (Wilcoxon *P* = 8.95×10⁻²¹), whereas eQTL CS sizes did not differ (p = 0.65; **Supplementary Figure 5d**), indicating that greater sQTL fine-mapping precision improves detection of shared causal variants. For loci classified as independent (PP.H3 > 0.8), most (67%) lacked CS overlap despite both traits being fine-mapped, further supporting distinct genetic mechanisms for splicing and expression regulation (**Supplementary Figure 5e**).

To characterize regulatory consequences of shared splicing-expression signals, we analyzed predicted coding outcomes of colocalized sQTL-eQTL pairs. At loci with strong evidence of colocalization, unproductive/NMD splicing was significantly more frequent than at independent (PP.H3) loci (**Fig. 4h, top**), and this enrichment persisted after stratifying by fine-mapped CS overlap (**Fig. 4h, bottom)**, implicating NMD as a mechanism linking splicing variation to expression at shared loci. Intersecting shared sQTL-eQTL signals with sQTL-GWAS colocalizations yielded 13 eQTL-sQTL-GWAS colocalizations across 9 GWAS loci; although limited in number, these cases represent high-confidence candidates for mechanistic follow-up, where splicing variation appears to influence both gene expression and disease susceptibility through shared regulatory mechanisms.

### Mechanistic insights from splicing-GWAS colocalization

One of the strongest PD colocalizations is a *cis*-sQTL at junction chr5:134,741,523–134,750,758 in calcium modulating ligand *CAMLG*. The sQTL (lead rs12657663) colocalizes with PD GWAS (PP.H4 = 0.97) and fine-maps to a single-variant 95% credible set with rs12657663 top (PIP = 1.00) (**Fig. 5a-b, top**). PD GWAS association (**Fig. 5b, bottom**) shows a concordant peak at the same position (−log_10_*P* ≈ 8) within the same narrow interval and LD relative to the PD index SNP rs11950533 indicates strong linkage (*r*² = 0.89) between rs12657663 and the GWAS peak, consistent with a shared causal signal (**Fig. 5a)**. Mechanistically, the sQTL promotes exon-3 skipping in *CAMLG*, a region overlapping predicted cytoplasmic, transmembrane, and intrinsically disordered segments (**Fig. 5c**), potentially affecting localization/anchoring or flexible interaction motifs. Leafcutter2’s^56^ classifies exon-3 skipping as productive (not NMD), and despite low baseline usage it shows a strong negative β that aligns with the negative GWAS β, suggesting the skipping allele may be protective. While *CAMLG* was previously implicated in PD by TWAS (DLPFC and monocytes)^54^, splicing was not proposed; moreover, the locus has been linked to PD-relevant lipidomic and lysosomal protein changes^57^. Together, these results highlight *CAMLG* as a compelling splicing-mediated effector gene contributing to PD susceptibility.

**Figure 5.**
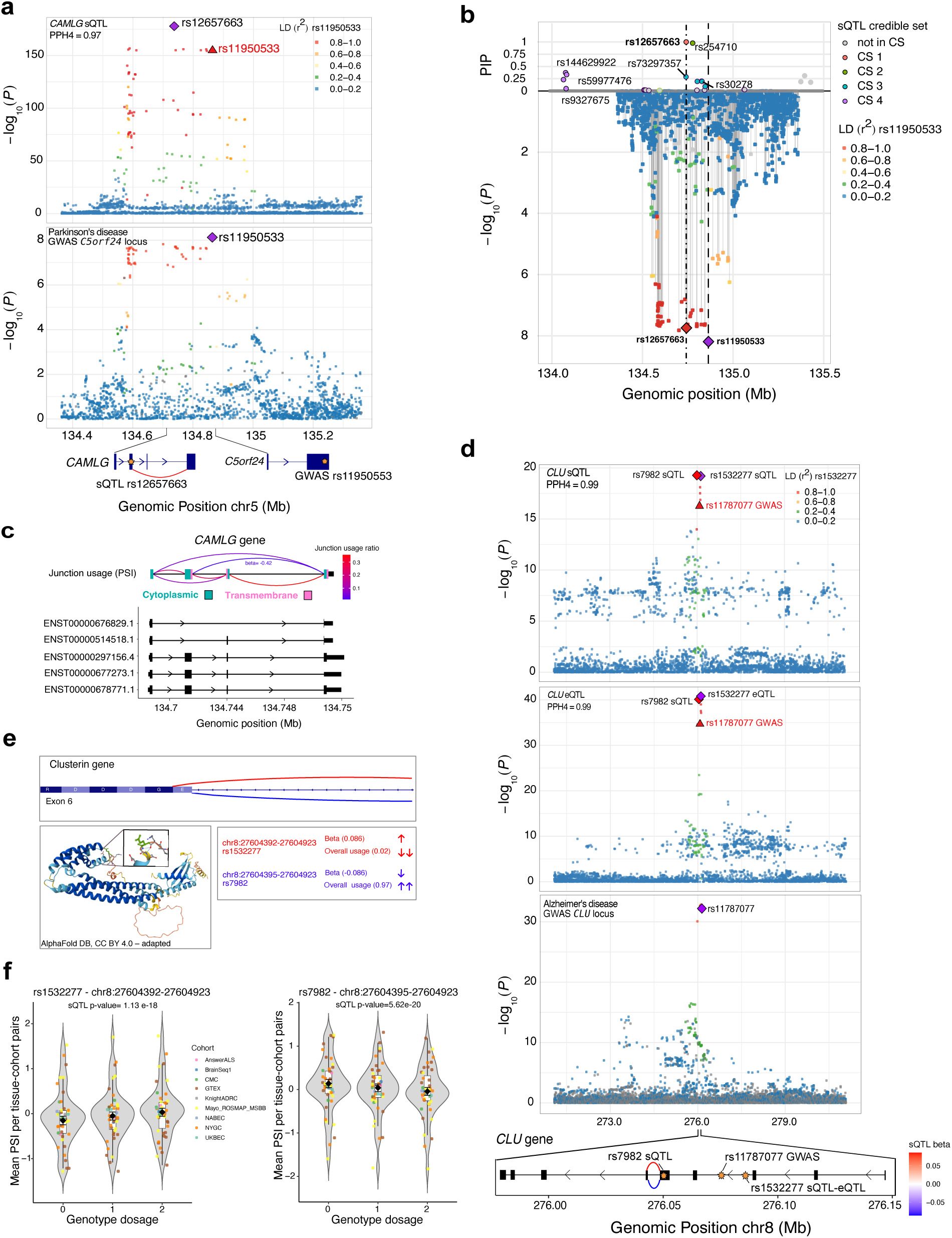
Example of two colocalizations of sQTLs with PD and AD GWAS signals. **(a)** Colocalization of *CAMLG* sQTL and Parkinson’s disease GWAS signals. Top: sQTL association −log₁₀(P) values for *CAMLG* junction; the fine-mapped sQTL variant rs12657663 is indicated (purple diamond). Bottom: GWAS −log₁₀(*P*) values for the Parkinson’s disease signal at the *C5orf24* locus; the GWAS lead SNP rs11950533 is marked (red triangle). LD is colored by r² with the sQTL lead SNP. **(b)** Mirror plot of sQTL fine-mapping and GWAS. Top: SuSiE PIP, dots color denotes 95% credible-set membership (CS1 red, CS2 green, CS3 blue, CS4 purple; open = not in CS). Bottom: single-variant sQTL −log₁₀(*P*). Each dot is a variant; fill encodes LD (r²) to AD GWAS index rs11950533. Diamonds highlight index variants (red = sQTL rs12657663, purple = GWAS rs11950533). Vertical dashed lines indicate their positions. **(c)** Schematic of *CAMLG* transcript structures and junction usage. Top: arcs represent splicing junctions colored by junction usage ratio (PSI). Middle: overlapping UniProt-annotated transmembrane and cytoplasmic domains. Bottom: Ensembl transcript models (hg38). **(d)** Colocalization of *CLU* sQTL and Alzheimer’s disease GWAS signals. Top: sQTL and eQTL association −log₁₀(*P*) values for *CLU* junctions; the fine-mapped sQTL variant rs1532277 is indicated (purple diamond). Bottom: GWAS −log₁₀(*P*) values for the Parkinson’s disease signal at the *CLU* locus; the GWAS lead SNP rs11787077 is marked (red triangle). LD is colored by *r*² with the sQTL lead SNP. **(e)** Schematic of *CLU* exon 6 showing a tandem 3′-acceptor (NAGNAG) pair spaced by 3 bp (positions chr8:27604392 and chr8:27604395). Splicing to the distal acceptor (395) deletes the first codon of exon 6, producing an in-frame Δ*E*; splicing to the proximal acceptor (392) retains that glutamate (*E*). Curved tracks summarize rs1532277 sQTL effect (β): red (β > 0) for the Δ*E* junction and blue (β < 0) for the E-retaining junction, indicating opposite allelic directions consistent with mutually exclusive acceptor usage. The AlphaFold model (pLDDT coloring) places the Δ*E* site in a low-confidence, flexible linker between α-helices (inset), suggesting minimal impact on the global fold and pointing instead to subtle changes in inter-helical spacing or isoform balance (bottom left panel). **(f)** Violin plot for the genotype-PSI relationships for rs1532277 and rs7982 across cohorts and tissues, shown separately for the two tandem-acceptor junctions. Each violin summarizes the distribution of per tissue-cohort mean PSI at each genotype dosage (0/1/2); boxes show median and IQR; points are individual tissue-cohort means; black diamonds mark the sample-size-weighted mean (± SE). Left (chr8:27604395–27604923-rs1532277, ΔE): PSI increases with risk-allele dosage (β > 0). Right (chr8:27604392–27604923-rs7982, E-retained): PSI decreases with risk-allele dosage (β < 0). AlphaFold prediction for human CLU (AFDB; EMBL-EBI & DeepMind), CC BY 4.0. Adapted (cropped/annotated).

Among the high number of sQTLs colocalizing with SCZ, we highlighted a *cis*-sQTL for junction chr8:17210480–17220255 in *ZDHHC2* which strongly colocalizes with SCZ GWAS (PP.H4 = 0.88; **Supplementary Figure 6a**). In the mirror plot (**Supplementary Figure 6b**; top = PIP, bottom = GWAS −log_10_*P*), fine-mapping concentrates sQTL probability into a narrow interval centered on rs13267258 (PIP = 0.49), and the GWAS peak aligns over the same region (LD to rs4921741 *r*² = 0.68). The affected junction skips the two exons immediately preceding the last exon (**Supplementary Figure 6c**), generating a frameshift and an unproductive isoform consistent with nonsense-mediated decay (NMD). Colocalization with eQTLs supports independent regulatory effects (PP.H3 = 1), consistent with transcript-level NMD altering RNA fate without a detectable change in bulk gene expression.

At *CLU* locus, we see multi-layer colocalization-splicing (sQTL) (**Fig. 5d top**), expression (eQTL) (**Fig. 5d, middle**), and AD GWAS (**Fig. 5d, bottom**). The AD lead rs11787077 colocalizes with both two sQTL and an eQTL; the sQTL involves two junctions, chr8:27604392–27604923 (rs1532277) and chr8:27604395–27604923 (rs7982), reflecting tandem NAGNAG 3′-acceptor usage in exon 6. One junction produces a 3-bp in-frame deletion removing a single glutamic acid (ΔE) (**Fig. 5e, top**). The junctions show opposite allelic directions and opposing AD associations. ΔE lies in a low-pLDDT linker between two α-helices in the AlphaFold model^58,59^, suggesting structural tolerance; effects likely arise from subtle changes in inter-helical spacing/interaction surfaces or isoform stability/trafficking, rather than gross disruption (**Fig. 5e, bottom**). Together with the eQTL, this supports a regulatory haplotype in which a small, tolerated splice shift (ΔE) accompanies *CLU* expression modulation. Genotype-PSI plots (**Fig. 5f**) show mirror-image dosage responses across cohorts-PSI increases with risk-allele dosage for the ΔE junction (chr8:27604395–27604923-rs1532277) and decreases for the E-retaining junction (chr8:27604392–27604923-rs7982); cohort-colored points cluster around sample-size-weighted means (black diamonds ± SE), indicating cross-dataset concordance.

## Discussion

Prior GWAS and transcriptomic studies have established AS as a fundamental molecular mechanism mediating disease risk^30,34,36,60^. However, the precise ways in which individual sQTLs influence transcript and protein isoform landscapes to mediate neurological traits are not fully characterized. In this study, we assembled BigBrain, a harmonized, population-scale map of splicing regulation in the human brain, consisting of 10,725 RNA-seq samples across 11 cohorts, multiple ancestries, and diverse brain regions. Our results reveal widespread genetic effects on alternative splicing, with the majority of genes having an sQTL and many having multiple independent sQTLs. Importantly, a substantial proportion of these sQTLs colocalize with neurological GWAS loci, underscoring the role of splicing variation in mediating genetic effects on disease susceptibility.

While several large-scale brain RNA-seq meta-analyses exist, most focus on gene-level regulatory effects, limiting mechanistic interpretation of genetic risk. BigBrain addresses this gap via splicing-resolved, fine-mapped brain QTLs with isoform-level mechanistic annotations to enable researchers to move from variant to isoform to functional consequence. This unlocks discovery in three ways: (i) it allows prioritization of causal variants, transcripts, and mechanisms at GWAS loci; (ii) it supports hypothesis generation for experimental follow-up, including protein and RNA regulatory consequences of splicing changes; and (iii) it provides data for benchmarking and training next-generation predictive models of genetic effects on splicing.

Although over half of sQTL credible sets overlapped with eQTLs, only a small subset of these jointly colocalized with GWAS loci. This suggests that splicing and expression capture complementary aspects of gene regulation, and that sQTLs mediate disease risk through mechanisms distinct from expression changes. As such, total expression analyses are insufficient to fully capture regulatory complexity, underscoring the need for isoform-level resolution and integrated interpretation of eQTL and sQTL data.

Despite these advances, important methodological challenges remain. Our approach enables the discovery of *cis*-regulatory splicing variation at population scale, but is limited to tissue resolution by bulk RNA-seq, which averages across heterogeneous cell populations. Many splicing events, particularly in the brain, are cell-type specific, and some isoforms are restricted to distinct neuronal or glial subtypes^61^. As such, regulatory effects may be diluted or obscured in bulk data, especially for rarer cell-types. Recent single-cell eQTL efforts (for example, our SingleBrain meta-analysis^62^ of ∼1,000 donors and 5.8M nuclei) show that cellular resolution can uncover regulatory variants missed in bulk and provide cell-type and even fine-grained cell-state context. However, extending these strategies to sQTL mapping is not feasible with existing population-scale single-nuclei RNA-seq datasets given their low read depth and extreme 3’ bias, restricting our ability to quantify splicing variation^63,64^.

At the same time, short-read RNA-seq limits our ability to resolve full-length isoforms and accurately interpret the downstream functional consequences of sQTLs. Our fine-mapping revealed multiple independent signals per gene, highlighting the complexity of splicing regulation, yet linking the impacted junctions to specific protein products remains challenging. Long-read RNA-seq could help disambiguate which isoforms are impacted by an sQTL, but to-date there are no population scale long-read RNA-seq postmortem brain datasets.

Even when the isoform-level change is clear, the downstream functional consequences may not be. For example, in the *CLU* locus, colocalized AD GWAS, sQTL, and eQTL signals converge on a junction differing by only 3 base pairs: a cryptic splice site would result in the loss of just one glutamic acid residue from the final protein product. This residue is not in a known protein domain, highlighting the need for functional experiments, such as CRISPR-based isoform-specific knockdowns^65^, to interpret the mechanistic consequences of sQTLs.

While BigBrain includes multiple ancestries, for which we provide full sQTL summary statistics, the integration with GWAS was performed using European-only sQTLs due to the limited GWAS in other ancestries. Expanding to global cohorts is crucial to capture ancestry-specific regulatory variation, enhance fine-mapping resolution through diverse LD structures, and ensure equitable biomedical insights. With the growing availability of multi-ancestry RNA-seq and GWAS data, there is an opportunity to uncover population-specific mechanisms of disease risk.

Together, these advances will refine our understanding of how genetic variants reshape isoform landscapes in a cell-type- and ancestry-specific manner, enabling splicing-aware therapeutic strategies, such as antisense oligonucleotides (ASOs), to restore or redirect splicing. BigBrain provides a foundational resource for splicing genetics in the brain and lays the groundwork for future studies integrating cell-type, isoform, and functional annotations to unravel the molecular mechanisms underlying genetic disease risk.

## Material and methods

Additional methods for analyses shared across BigBrain Project meta-analysis are provided in the **Supplementary Note**.

RNA-sequencing and matched genotype data were collected from 11 publicly available brain tissue cohorts, totaling 10,725 samples from 4,656 unique individuals (**Table 1**). Both expression and genotype data were uniformly processed and harmonized across cohorts in the BigBrain project.

### Genomic Sequencing Quality Control

Genotype QC was performed using an in-house PLINK-based pipeline^66^. For SNP arrays, variants were filtered using bcftools v.1.9 and VCFtools v.0.1.15, removing those with a call rate below 95%, a minor allele frequency (MAF) below 5%, and a Hardy-Weinberg equilibrium p-value below 1 × 10⁻⁶. Imputation was conducted using the Michigan Imputation Server^67^ with the 1000 Genomes Phase 3 European reference panel^68^, and variants were lifted from GRCh37 to GRCh38 post-imputation, in order to match the RNA-seq samples. A final QC step removed indels, multiallelic SNPs, and retained SNPs with MAF > 5% and HWE P > 1e-6. Whole-genome sequencing (WGS) data underwent Variant Quality Score Recalibration (VQSR) with GATK, applying stringent variant- and sample-level filters (See **Supplementary Note**). To ensure consistency across datasets, RNA-seq samples were matched to donors using MBV^69^ (QTLtools v1.3), genetic ancestry inferred by Somalier^70^, and samples with duplicated or third-degree kinship (KING v2.2.5)^69^ were removed. Final VCFs excluded singletons and harmonized SNP IDs across cohorts (See **Supplementary Note, Table 2**).

### Junction quantification

RNA-seq data across cohorts varied in library preparation protocols, including differences in strandedness, read length, and sequencing depth. Given the scale and diversity of the dataset, including 43 tissue-cohort pairs across multiple brain regions and populations, such protocol variability posed a significant challenge for consistent splicing quantification. While we did not harmonize these parameters explicitly during alignment or junction extraction, we addressed their downstream impact using normalization and covariate modeling. After alignment to the reference genome, junction counts were derived from RNA-seq data in each cohort using Leafcutter^43^ (See Pre-processing RNA-sequencing data and Mapping RNA-sequencing data in **Supplementary Note**). Junctions were extracted from BAM files using the regtools software (regtools.readthedocs.org). The *junctions extract* function from regtools leverages Compact Idiosyncratic Gapped Alignment Report (CIGAR) strings from BAM files to accurately quantify the usage of each intron, providing a reliable and streamlined method for performing splicing analyses. Junctions expressed in 50% or more of the samples per cohort were retained. Across the 43 tissue-cohort pairs, junctions shared by at least 12 pairs were kept for intron clustering. Intron clustering was defined using LeafCutter^43^ globally across all junctions in all tissue-cohorts to harmonize splicing clusters across datasets. This process clusters introns extracted by regtools, requiring at least 50 split reads to support each cluster.

Percent Spliced In (PSI) values were calculated for each junction based on the proportion of spliced reads supporting the junction relative to the total spliced reads within the associated cluster. Junctions missing in more than 50% of samples in every tissue-cohort pair were removed, along with those with PSI values of 0 or 1 in 90% of samples. Remaining missing values were imputed with the median junction PSI within each tissue-cohort pair, resulting in a total of 81,960 junctions shared across tissue-cohorts. Resulting PSI matrices were scaled, centered, and quantile-normalized for each tissue-cohort pair to minimize technical and cohort-specific biases.

### Principal component analysis

PCA was performed on both raw imputed and normalized PSI matrices using the irlba package (https://github.com/bwlewis/irlba), retaining 50 PCs. PSI for 81,970 junctions that passed the final filtering step was logit-transformed, with a pseudocount of 0.01 applied to each junction’s PSI before PCA. To evaluate the influence of metadata variables on splicing variation, linear regression models were applied to each principal component (PC) using cohort, brain region and as predictors. The proportion of variance explained by these variables was assessed using adjusted R² values, indicating the strength of association between each PC and metadata.

### sQTL mapping and meta-analysis with mmQTL

Multi-ancestry mixed model (mmQTL) (https://github.com/jxzb1988/MMQTL/tree/v1.5.0^42^) was used for junction-QTL mapping (sQTL) meta-analysis. mmQTL enhances statistical power by aggregating associations across tissue-cohort pairs while accounting for effect-size heterogeneity, population structure and relatedness, and sample overlap/repeated donors^42^. Normalized junction PSI matrices were adjusted for 15 PEER factors, and age and sex (inferred using Somalier) were included as covariates. QTL mapping was performed with mmQTL v.26a using a linear mixed model, followed by a random-effects meta-analysis across 43 datasets. Genotypes were converted to PLINK v.2.3 format, and a relatedness matrix was generated with GCTA v.1.94.1. SNPs (MAF > 0.01) within 1MB windows were tested. Random-effects p-values were Bonferroni-corrected per feature and further subjected to Benjamini–Hochberg FDR correction across all features. (See extended Material and Methods for more details). The correlation of effect sizes of sQTL between tissue-cohort pairs included in the mmQTL meta-analysis was computed using a Pearson pairwise correlation, highlighting per tissue and per cohort correlation examples. Distance from acceptor (end) and donor (start) site of the splice junction was also calculated for significant sQTL from mmQTL meta-analysis in a strand specific approach. To quantify discovery rates across individual cohorts and the meta-analysis, Z-scores for each tested junction were extracted and converted into two-sided p-values. A Bonferroni correction was applied to adjust for multiple testing, with significance defined as P < 0.05 divided by the number of tested junctions. For each cohort, we computed the number of significantly associated junctions and the total number tested. The discovery rate was calculated as the proportion of significant associations among all tested junctions. Cohort-level summaries were integrated with study and sample size annotations for comparative evaluation.

### sQTL replication analysis

To assess the replicability of the mmQTL sQTL associations across all tissue-cohort pairs, an mmQTL meta-analysis excluding ROSMAP-DPLCF1 as the held-out cohort was performed. Significant associations were then tested in the held-out cohort using the qvalue package (https://github.com/StoreyLab/qvalue), with P values adjusted using the Benjamini-Hochberg FDR ^71^. Additionally, sQTL analysis on the ROSMAP-DPLCF1 cohort alone was performed and these associations were tested in the remaining cohorts from the meta-analysis. Associations were considered replicable if they met FDR < 0.05 and showed the same effect direction in both the meta-analysis (excluding the held-out cohort) and the ROSMAP-DPLCF1 cohort.

### Fine mapping

PolyFun^26^ (https://github.com/omerwe/polyfun) was used for functionally informed fine-mapping and used SuSiE^47,48^ (https://github.com/stephenslab/susieR) as the fine-mapping method.

#### Summary Statistics (SNPs extract and munge)

Junctions with top sQTLs were extracted from the full summary statistics generated by mmQTL mapping. SNP summary statistics were processed using the munge_polyfun_sumstats.py script from the PolyFun package^26^, modified to exclude variants zero or missing across all tissue-cohort pairs. This step removed variants missing across all tested tissue-cohort but kept some missingness and variability. To address differences in variance across tissues, random Z-scores were used in place of fixed Z-scores, aligning their signs to those of the original fixed Z-scores.

#### Genotype Data and LD Matrix

Individual genotype data from 3,884 unique participants of European ancestry in the BigBrain study was used to compute the in-sample linkage disequilibrium (LD) matrix. This ensured consistency between the LD matrix and the genotype data used to derive sQTL statistics. From the 9,130,833 SNPs tested across all tissue-cohort pairs in the sQTL analysis, we selected SNPs present in at least 21 tissue-cohort pairs (51% of all pairs), resulting in 7,375,120 highly shared SNPs (81% of the total) for the LD matrix calculation. Genotype data from VCF files across multiple cohorts were converted into PLINK format (.bed, .bim, and .fam) using PLINK package (http://pngu.mgh.harvard.edu/purcell/plink/v1.9). LD matrices were calculated separately for each chromosome to enhance computational efficiency.

#### Fine-Mapping

Fine-mapping was conducted using the finemapper.py script from the PolyFun package^26^ (https://github.com/omerwe/polyfun) in a 1 Mb window around both the start and the end positions of the junctions. SuSiE was conFig.d with a maximum of 5 causal variants per locus, and the following options were applied: --no-sort-pip: Prevent sorting results by PIP values; --allow-swapped-indel-alleles: Retain and process indels with swapped alleles between summary statistics and the LD matrix; --susie-resvar 1: Use a fixed residual variance value. The analysis was performed per chromosome for computational efficiency. Fine-mapping results were later combined across chromosomes for downstream analysis.

To evaluate the genomic localization of fine-mapped sQTLs, SNP coordinates were intersected with curated BED annotations for coding exons, introns, splice sites, untranslated regions (UTRs), promoters, enhancers, and RNA-binding protein (RBP) sites using the GenomicRanges::findOverlaps function. Enrichment was assessed by comparing the proportion of fine-mapped SNPs (posterior inclusion probability, PIP > 0.5) overlapping each category to a background set of 75,000 SNPs with PIP ≈ 0. Statistical significance was determined using Fisher’s exact test, and multiple comparisons were corrected using the Benjamini-Hochberg procedure. To assess positional biases, distances were calculated from each fine-mapped SNP to the nearest donor and acceptor splice site (strand-aware), as well as to the midpoint of the associated junction or gene body. For selected loci of interest, we visualized SuSiE fine-mapping results by plotting posterior inclusion probabilities (PIPs) for all variants within the tested locus. We also compared the distribution of log₁₀ Bayes Factors (logBF) across PIP thresholds to evaluate the strength of fine-mapped signals.

### Protein domain analysis

leafidex R package (https://github.com/atokolyi/leafidex) was used to map splicing junctions identified by Leafcutter^43^ to protein domains from the UniProt and Pfam databases using a 10bp expanded window from both start and end. In addition to the original annotations provided by the package, its functionality was extended by incorporating the `unipInterest` module from UniProt, which captures functionally relevant protein regions. Furthermore, a list of DNA- and RNA-binding protein domains was manually curated by extracting domain-level annotations from UniProt. These annotations were derived using the output of TappAS (https://github.com/ConesaLab/tappAS), run on GENCODE v39 transcript models. This allowed us to integrate high-confidence functional regions associated with nucleic acid binding into our splicing analysis. The junction-gene and protein mapping was performed in the total 81,960 junctions tested for sQTL mapping and significant sQTL were subsequently subgroups for following analysis and on the 116,099 constitutive junctions defined as the junctions with PSI ≤ 1% were within their cluster and not tested for sQTL analysis.

To classify sQTL in quantile bins based on effect size (beta) we used the |beta| obtained running sQTL mapping per tissues-cohort pairs independently using tensorQTL^72^ on junctions quantification prior scaled/center and quantile normalization and without any PEER correction to avoid inflated effect derived for normalization steps. For all significant sQTLs in the meta-analysis, we took the absolute value |β| for the tissue-cohort pair runs and binned into quartiles based on their distribution in our dataset. Non-significant tests were retained as the non-sQTL group (with |β| set to 0 for analyses that require an effect size). Unless stated otherwise, downstream summaries aggregate results across tissue-cohort runs while preserving these within-run bin labels.

The GeneBayes framework for gene constraint (s_het_) estimation from Zeng et al.^51^ were used as a measure of gene constraint. Genes were stratified into four quantile-based bins (Q1-Q4) and grouped into broader categories: low constraint (Q1-Q2), moderate constraint (Q3), and high constraint (Q4) for downstream analysis on protein domains. Junctions were further classified according to their mapped GeneBayes classification from Zeng et al.^51^ for downstream analysis on protein domains.

Association of domain localization and gene constraint with sQTLs. Using junctions with complete annotation, we first ran unadjusted Pearson χ² tests within each GeneBayes constraint tier (Low/Moderate/High) to assess association between sQTL status (sQTL vs non-sQTL) and protein-domain class, and analogously for the effect-size outcome (high-|β| vs low; non-sQTLs set to β=0, high defined as the |β| ≥ 90th percentile corresponding to beta threshold =0.015). When expected counts were <5 we used Fisher’s exact test; effect size was summarized by Cramér’s V; P values were BH-adjusted within outcome. We then fit two sets of logistic models. (i) For sQTL presence, among CDS-overlapping junctions we modeled sQTL status as a function of domain class, GeneBayes constraint group, their interaction, and total domain overlap (covariate). (ii) For effect size, high-effect sQTLs were defined as the |β| ≥ 90th percentile corresponding to beta threshold = 0.015 (non-sQTLs assigned β=0), and we modeled high vs low |β| with the same predictors. For each domain we evaluated domain-only, constraint-only, and domain×constraint interaction models, fitted with glm(family=binomial) in R; results were obtained with broom::tidy() and Benjamini–Hochberg FDR was applied across tests.

### Frameshift classification and NMD potential assessment

Junctions overlapping protein-coding regions were analyzed to assess potential frameshift-inducing effects. Junctions were first filtered to retain only those annotated as overlapping coding sequences (CDS) using leafidex domain-level annotations. Splice junctions from regtools (BED; 0-based, half-open) were converted to 1-based, closed coordinates by adding +1 to starts. Coding sequence (CDS) intervals were from GENCODE v46 (GTF; 1-based, closed). Junction spans were intersected with CDS exons in a strand-specific manner using bedtools intersect -s -wa -wb. For each junction-CDS pair we computed inclusive base-pair overlap and flagged boundary adjacency when the intron abutted, but did not overlap, a CDS edge (1-based closed adjacency: intron start = exon end + 1, or intron end = exon start − 1). Pairs with positive overlap or boundary adjacency were retained.

For each junction-transcript combination we then applied a two-part rule: a) Overlap rule. We summed CDS overlap across that transcript’s CDS exons. If total overlap > 0 and divisible by 3 we labeled in-frame (overlap); if > 0 and not divisible by 3 we labeled frameshift (overlap). b) Boundary rule (no overlap). When overlap = 0 but boundary adjacency existed, we distinguished: a) Both-touch (exon skipping): if the intron touched a CDS exon at both ends, we inferred the skipped CDS length as the distance between the nearest CDS end upstream of the junction start and the nearest CDS start downstream of the junction end (taken from the CDS boundaries present in the intersection). If the skipped length mod 3 = 0 → in-frame (both-touch); otherwise → frameshift (both-touch). b) One-touch: if exactly one end touched a CDS edge, we measured the distance from the non-touching intron end to the next CDS splice boundary across the intron (again using only CDS boundaries present in the intersection). If this distance mod 3 = 0 → in-frame (one-touch); otherwise → frameshift (one-touch). c) If the partner boundary could not be located within CDS (e.g., partner lies in UTR and was not present in the CDS set), the case was kept as boundary_only (unresolved). Per-transcript calls were then collapsed to one label per junction: ambiguous only when the junction had both in-frame and frameshift evidence across transcripts; otherwise in-frame or frameshift according to the available evidence. Non-CDS and boundary_only evidence did not contribute to ambiguity (junctions with only boundary_only evidence were labeled boundary_only). Proportions were reported as the fraction of junctions in each final category.

Splice junctions from *regtools* output for each sample were re-clustered using the leafCutter2 package (https://github.com/cfbuenabadn/leafcutter2/tree/main)^56^, which enables functional annotation of junctions as *productive* (PR), *unproductive* (UP), *intron retention* (IN), or *ambiguous/NE*. leafCutter2 incorporates transcript models and strand-specific coding sequence (CDS) boundaries to infer the likelihood of nonsense-mediated decay (NMD) or productive protein-coding outcomes to systematically evaluate the impact of sQTLs on transcript productivity. To evaluate differences in coding consequences between in-frame and frameshift-inducing sQTL junctions, two-by-two Fisher’s exact tests were performed separately for each unproductive status category (PR, UP, NE, IN). For each category, the observed counts of frameshift and in-frame events were compared against the remaining junctions. P-values were adjusted for multiple testing using the Benjamini-Hochberg false discovery rate (FDR) correction. Analyses were conducted in R using the fisher.test() function, and adjusted p-values were calculated with p.adjust (method = “fdr”).

### Colocalization analysis

COLOC^55^ was used to test SNP colocalization between GWAS for Alzheimer’s disease (AD), ALS, bipolar disorder (BPD), Parkinson’s disease (PD), schizophrenia (SCZ), Major depressive disorder (MDD), Multiple sclerosis (MS) and Rheumatoid arthritis (RA) and sQTLs. The analysis was restricted to sQTL results for European individuals only and focused on SNPs within 1MB of significant loci, with LD assessed using the 1000 Genomes European populations. Colocalizations were considered for GWAS-QTL SNP pairs that met the following criteria:(i) the SNP had a GWAS p-value < 5×10⁻⁸; (ii) the posterior probability of a shared causal variant (PP.H4) was ≥ 0.8; (iii) distance filters were applied as follows: (a) the GWAS and QTL SNPs were within 100 kb of each other; (b) the GWAS SNP and the center of the associated junction were within 100 kb; (c) the QTL SNP and the center of the associated junction were within 100 kb, or the GWAS and QTL SNPs were within 500 kb and had LD > 0.1. Full details, including filtering criteria and methodology, are provided in the extended materials (Extended Materials and Methods). To additionally capture high-confidence colocalizations driven by fine-mapped causal variants, we incorporated an orthogonal filter based on overlap with 95% sQTL credible sets. Specifically, we intersected all GWAS summary statistics with sQTL credible set variants and retained any GWAS-sQTL SNP pair showing overlap, provided it also satisfied the main statistical thresholds (GWAS p-value < 5×10⁻⁸ and PP.H4.abf ≥ 0.8), regardless of physical proximity.

### eQTL-sQTL colocalization analysis

Colocalization analysis between sQTLs and eQTLs was performed using COLOC^55^, applying the same probabilistic model used in the sQTL-GWAS analysis. The analysis was restricted to individuals of European ancestry and included all SNPs within ±1 Mb of tested splice junctions and eGenes, without additional filtering. A posterior probability (PP.H4) ≥ 0.8 was used to identify loci where the genetic signal was shared between sQTLs and eQTLs. Independent effects were defined by PP.H3 ≥ 0.8. Trait-specific associations were identified using PP.H1 ≥ 0.8 for eQTLs and PP.H2 ≥ 0.8 for sQTLs. No further p-value or distance-based filtering was applied. Full methodological details are available in the Extended Materials and Methods.

To assess whether colocalized eQTL-sQTL signals may be influenced by the same underlying genetic signal, the overlap between 95% credible sets from SuSiE fine-mapping for each trait was examined. SNPs not assigned to any credible set (CS = 0) and with PIP < 0.5 were filtered out. SNPs with CS = 0 but PIP ≥ 0.5 were retained, although none were present in our dataset. For each gene-junction pair with strong colocalization (PP.H4 ≥ 0.8), shared SNPs between the sQTL and eQTL fine-mapping results were identified. Pairs with at least one overlapping SNP were labeled as “shared CS (SNP overlap).” Remaining colocalized pairs without overlapping SNPs were further categorized based on whether both the eQTL and sQTL signals had been successfully fine-mapped. These were classified as: (1) “No overlap but both fine-mapped,” (2) “eQTL not fine-mapped,” (3) “sQTL not fine-mapped,” or (4) “Neither fine-mapped.” Additionally, lead SNP matching was assessed for SNP in shared CS. The same analysis was performed on eQTL-sQTL independent signals (H3 >0.8).

To assess potential functional effects, we annotated sQTL-associated junctions with transcript productivity status. Each junction was classified as productive (PR), unproductive (UP) due to predicted nonsense-mediated decay, non-coding (IN), or ambiguous effect (NE) based on transcript structure and open reading frame integrity. These annotations were used to compare the distribution of junction functional classes across fine-mapping colocalization categories, and enrichment was evaluated using Fisher’s exact tests.

## Statistics & Reproducibility

No statistical methods were used to predefine sample size. All available data were included in the analyses, and no exclusions were made. Experiments were not randomized, and investigators were not blinded to group allocation or outcome assessment. For all QTL analyses, data were assumed to follow a normal distribution after log-normal transformation, though this assumption was not formally tested.

## Acknowledgements

We thank the patients and families for their generous gift of brain donation. We are thankful to Gloria Sheynkman for helpful feedback and discussions.

This study was supported by the following National Institutes of Health grants:

NIA U01-AG068880, NIA R21-AG063130, NIA R01-AG054005, NIA RF1-AG065926, NIA R01-AG065926 NIA R56-AG088669, NIA R21-AG091272, NIA P30-AG066514, NINDS U54-NS123743, and NINDS R01-NS116006 to TR, JH, BZM, KBP, BJ, and WHC; NIA U01-AG068880 to TR, JH, BZM, KBP, BJ, WHC, AR, and DAK.

This work was supported in part through the computational resources and staff expertise provided by Scientific Computing at the Icahn School of Medicine at Mount Sinai and supported by the Clinical and Translational Science Awards (CTSA) grant UL1TR004419 from the National Center for Advancing Translational Sciences. Research reported in this paper was supported by the Office of Research Infrastructure of the National Institutes of Health under award number S10OD026880 and S10OD030463. The content is solely the responsibility of the authors and does not necessarily represent the official views of the National Institutes of Health.

## Data availability

No sequencing data was generated as part of this project. Data accession numbers for each cohort are listed in the **Supplementary Note**. All summary statistics from QTL analyses are made publicly available on our portal (https://bigbrain.nygenome.org/) and on Zenodo 10.5281/zenodo.17153730 (sQTL), 10.5281/zenodo.17226890 (eQTL), 10.5281/zenodo.17210092 (edQTL), 10.5281/zenodo.17297440 (QTL-QTL colocalization).

## Code Availability

Genotype quality control pipeline: https://github.com/RajLabMSSM/Genotype_QC_Pipeline_2.0 QTL preparation and meta-analysis pipeline: https://github.com/RajLabMSSM/mmQTL-pipeline Colocalization and MESC pipelines: 10.5281/zenodo.13864729, https://github.com/RajLabMSSM/downstream-QTL

## Author Contributions

The study was conceived by DAK, JH, and TR. Data analysis was performed by AR, with contributions from KBP, WHC, JH, BZM, BJ, HHW, TR and DAK. AT constructed the website. The work was supervised by DAK and TR. The manuscript was written by AR and revised by DAK, with input from all co-authors. All authors have read and approved the final manuscript.

## References

1. Nilsen, T. W. & Graveley, B. R. Expansion of the eukaryotic proteome by alternative splicing. Nature 463, 457–463 (2010).

2. Fair, B. et al. Global impact of unproductive splicing on human gene expression. Nat. Genet. 56, 1851–1861 (2024).

3. Wang, E. T. et al. Alternative isoform regulation in human tissue transcriptomes. Nature 456, 470–476 (2008).

4. Barbosa-Morais, N. L. et al. The evolutionary landscape of alternative splicing in vertebrate species. Science 338, 1587–1593 (2012).

5. Merkin, J., Russell, C., Chen, P. & Burge, C. B. Evolutionary dynamics of gene and isoform regulation in Mammalian tissues. Science 338, 1593–1599 (2012).

6. Licatalosi, D. D. & Darnell, R. B. Splicing regulation in neurologic disease. Neuron 52, 93–101 (2006).

7. Parikshak, N. N. et al. Genome-wide changes in lncRNA, splicing, and regional gene expression patterns in autism. Nature 540, 423–427 (2016).

8. Lenzken, S. C., Achsel, T., Carrì, M. T. & Barabino, S. M. L. Neuronal RNA-binding proteins in health and disease. Wiley Interdiscip. Rev. RNA 5, 565–576 (2014).

9. International Multiple Sclerosis Genetics Consortium. Multiple sclerosis genomic map implicates peripheral immune cells and microglia in susceptibility. Science 365, eaav7188 (2019).

10. Wray, N. R. et al. Genome-wide association analyses identify 44 risk variants and refine the genetic architecture of major depression. Nat. Genet. 50, 668–681 (2018).

11. Nicolas, A. et al. Genome-wide analyses identify KIF5A as a novel ALS gene. Neuron 97, 1267–1288 (2018).

12. van Rheenen, W. et al. Common and rare variant association analyses in amyotrophic lateral sclerosis identify 15 risk loci with distinct genetic architectures and neuron-specific biology. Nat. Genet. 53, 1636–1648 (2021).

13. Trubetskoy, V. et al. Mapping genomic loci implicates genes and synaptic biology in schizophrenia. Nature 604, 502–508 (2022).

14. Bellenguez, C. et al. New insights into the genetic etiology of Alzheimer’s disease and related dementias. Nat. Genet. 54, 412–436 (2022).

15. Lambert, J. C. et al. Meta-analysis of 74,046 individuals identifies 11 new susceptibility loci for Alzheimer’s disease. Nat. Genet. 45, 1452–1458 (2013).

16. Marioni, R. E. et al. GWAS on family history of Alzheimer’s disease. Transl. Psychiatry 8, (2018).

17. Jansen, I. E. et al. Genome-wide meta-analysis identifies new loci and functional pathways influencing Alzheimer’s disease risk. Nat. Genet. 51, 404–413 (2019).

18. Kunkle, B. W. et al. Genetic meta-analysis of diagnosed Alzheimer’s disease identifies new risk loci and implicates Aβ, tau, immunity and lipid processing. Nat. Genet. 51, 414–430 (2019).

19. Nalls, M. A. et al. Identification of novel risk loci, causal insights, and heritable risk for Parkinson’s disease: a meta-analysis of genome-wide association studies. Lancet Neurol. 18, 1091–1102 (2019).

20. Schizophrenia Working Group of the Psychiatric Genomics Consortium. Biological insights from 108 schizophrenia-associated genetic loci. Nature 511, 421–427 (2014).

21. Mullins, N. et al. Genome-wide association study of more than 40,000 bipolar disorder cases provides new insights into the underlying biology. Nat. Genet. 53, 817–829 (2021).

22. Gasperini, M., Tome, J. M. & Shendure, J. Towards a comprehensive catalogue of validated and target-linked human enhancers. Nat. Rev. Genet. 21, 292–310 (2020).

23. Peña-Martínez, E. G. & Rodríguez-Martínez, J. A. Decoding non-coding variants: Recent approaches to studying their role in gene regulation and human diseases. Front. Biosci. (Schol. Ed.) 16, 4 (2024).

24. Ying, P. et al. Genome-wide enhancer-gene regulatory maps link causal variants to target genes underlying human cancer risk. Nat. Commun. 14, 5958 (2023).

25. Schaid, D. J., Chen, W. & Larson, N. B. From genome-wide associations to candidate causal variants by statistical fine-mapping. Nat. Rev. Genet. 19, 491–504 (2018).

26. Weissbrod, O. et al. Functionally informed fine-mapping and polygenic localization of complex trait heritability. Nat. Genet. 52, 1355–1363 (2020).

27. GTEx Consortium et al. Genetic effects on gene expression across human tissues. Nature 550, 204–213 (2017).

28. GTEx Consortium. Human genomics. The Genotype-Tissue Expression (GTEx) pilot analysis: multitissue gene regulation in humans. Science 348, 648–660 (2015).

29. Dimas, A. S. et al. Common regulatory variation impacts gene expression in a cell type-dependent manner. Science 325, 1246–1250 (2009).

30. GTEx Consortium. The GTEx Consortium atlas of genetic regulatory effects across human tissues. Science 369, 1318–1330 (2020).

31. Barbeira, A. N. et al. Exploiting the GTEx resources to decipher the mechanisms at GWAS loci. Genome Biol. 22, 49 (2021).

32. Yao, D. W., O’Connor, L. J., Price, A. L. & Gusev, A. Quantifying genetic effects on disease mediated by assayed gene expression levels. Nat. Genet. 52, 626–633 (2020).

33. Chun, S. et al. Limited statistical evidence for shared genetic effects of eQTLs and autoimmune-disease-associated loci in three major immune-cell types. Nat. Genet. 49, 600–605 (2017).

34. Li, Y. I. et al. RNA splicing is a primary link between genetic variation and disease. Science 352, 600–604 (2016).

35. Garrido-Martín, D., Borsari, B., Calvo, M., Reverter, F. & Guigó, R. Identification and analysis of splicing quantitative trait loci across multiple tissues in the human genome. Nat. Commun. 12, 727 (2021).

36. Qi, T. et al. Genetic control of RNA splicing and its distinct role in complex trait variation. Nat. Genet. 54, 1355–1363 (2022).

37. El Garwany, O., et al. Low-usage splice junctions underpin immune-mediated disease risk. bioRxiv (2023) doi:10.1101/2023.05.29.542728.

38. Wen, C. et al. Cross-ancestry atlas of gene, isoform, and splicing regulation in the developing human brain. Science 384, eadh0829 (2024).

39. Walker, R. L. et al. Genetic control of expression and splicing in developing human brain informs disease mechanisms. Cell 181, 745 (2020).

40. Hormozdiari, F. et al. Leveraging molecular quantitative trait loci to understand the genetic architecture of diseases and complex traits. Nat. Genet. 50, 1041–1047 (2018).

41. Raj, T. et al. CD33: increased inclusion of exon 2 implicates the Ig V-set domain in Alzheimer’s disease susceptibility. Hum. Mol. Genet. 23, 2729–2736 (2014).

42. Zeng, B. et al. Multi-ancestry eQTL meta-analysis of human brain identifies candidate causal variants for brain-related traits. Nat. Genet. 54, 161–169 (2022).

43. Li, Y. I. et al. Annotation-free quantification of RNA splicing using LeafCutter. Nat. Genet. 50, 151–158 (2018).

44. Stegle, O., Parts, L., Piipari, M., Winn, J. & Durbin, R. Using probabilistic estimation of expression residuals (PEER) to obtain increased power and interpretability of gene expression analyses. Nat. Protoc. 7, 500–507 (2012).

45. Dredge, W. H. et al. Meta-analysis of genetic regulation of RNA editing in the human brain identifies new genes underlying neurological disease. medRxiv (2025) doi:10.1101/2025.09.30.25337026.

46. Zhang, Y. et al. Regional variation of splicing QTLs in human brain. Am. J. Hum. Genet. 107, 196–210 (2020).

47. Zou, Y., Carbonetto, P., Wang, G. & Stephens, M. Fine-mapping from summary data with the ‘Sum of Single Effects’ model. PLoS Genet. 18, e1010299 (2022).

48. Wang, G., Sarkar, A., Carbonetto, P. & Stephens, M. A simple new approach to variable selection in regression, with application to genetic fine mapping. J. R. Stat. Soc. Series B Stat. Methodol. 82, 1273–1300 (2020).

49. Agirre, E., Oldfield, A. J., Bellora, N., Segelle, A. & Luco, R. F. Splicing-associated chromatin signatures: a combinatorial and position-dependent role for histone marks in splicing definition. Nat. Commun. 12, 682 (2021).

50. Yustis, J.-C., Devoucoux, M. & Côté, J. The functional relationship between RNA splicing and the chromatin landscape. J. Mol. Biol. 436, 168614 (2024).

51. Zeng, T., Spence, J. P., Mostafavi, H. & Pritchard, J. K. Bayesian estimation of gene constraint from an evolutionary model with gene features. Nat. Genet. 56, 1632–1643 (2024).

52. Glassberg, E. C., Gao, Z., Harpak, A., Lan, X. & Pritchard, J. K. Evidence for weak selective constraint on human gene expression. Genetics 211, 757–772 (2019).

53. Stahl, E. A. et al. Genome-wide association study identifies 30 loci associated with bipolar disorder. Nat. Genet. 51, 793–803 (2019).

54. Li, Y. I., Wong, G., Humphrey, J. & Raj, T. Prioritizing Parkinson’s disease genes using population-scale transcriptomic data. Nat. Commun. 10, 994 (2019).

55. Wallace, C. Statistical testing of shared genetic control for potentially related traits. Genet. Epidemiol. 37, 802–813 (2013).

56. Najar, C. F. B. A. et al. Genetic and functional analysis of unproductive splicing using LeafCutter2. bioRxivorg (2025) doi:10.1101/2025.04.06.646893.

57. Zhang, H. et al. Calcium modulating ligand confers risk for Parkinson’s disease and impacts lysosomes. Ann. Clin. Transl. Neurol. 12, 925–937 (2025).

58. Jumper, J. et al. Highly accurate protein structure prediction with AlphaFold. Nature 596, 583–589 (2021).

59. Fleming, J. et al. AlphaFold Protein Structure Database and 3D-Beacons: New data and capabilities. J. Mol. Biol. 437, 168967 (2025).

60. Yamaguchi, K. et al. Splicing QTL analysis focusing on coding sequences reveals mechanisms for disease susceptibility loci. Nat. Commun. 13, 4659 (2022).

61. Joglekar, A. et al. A spatially resolved brain region- and cell type-specific isoform atlas of the postnatal mouse brain. Nat. Commun. 12, 463 (2021).

62. Jang, B. et al. SingleBrain: A meta-analysis of single-nucleus eQTLs linking genetic risk to brain disorders. medRxiv (2025) doi:10.1101/2025.03.06.25323424.

63. Gupta, I. et al. Single-cell isoform RNA sequencing characterizes isoforms in thousands of cerebellar cells. Nat. Biotechnol. 36, 1197–1202 (2018).

64. Arzalluz-Luque, Á. & Conesa, A. Single-cell RNAseq for the study of isoforms-how is that possible? Genome Biol. 19, 110 (2018).

65. Schertzer, M. D. et al. Cas13d-mediated isoform-specific RNA knockdown with a unified computational and experimental toolbox. Nat. Commun. 16, 6948 (2025).

66. Purcell, S. et al. PLINK: a tool set for whole-genome association and population-based linkage analyses. Am. J. Hum. Genet. 81, 559–575 (2007).

67. Das, S. et al. Next-generation genotype imputation service and methods. Nat. Genet. 48, 1284–1287 (2016).

68. 1000 Genomes Project Consortium et al. A global reference for human genetic variation. Nature 526, 68–74 (2015).

69. Manichaikul, A. et al. Robust relationship inference in genome-wide association studies. Bioinformatics 26, 2867–2873 (2010).

70. Pedersen, B. S. et al. Somalier: rapid relatedness estimation for cancer and germline studies using efficient genome sketches. Genome Med. 12, 62 (2020).

71. Benjamini, Y. & Hochberg, Y. Controlling the false discovery rate: A practical and powerful approach to multiple testing. J. R. Stat. Soc. Series B Stat. Methodol. 57, 289–300 (1995).

72. Taylor-Weiner, A. et al. Scaling computational genomics to millions of individuals with GPUs. Genome Biol. 20, 228 (2019).

