## Supplementary Notes for "Mapping genetic effects on splicing in ten thousand post-mortem brain samples reveals novel mediators of neurological disease risk"

### Supplementary Methods

### Transcriptomic datasets included in the BigBrain Project

I. Answer ALS
   A. The Answer ALS Research study includes subjects with sporadic or familial amyotrophic lateral sclerosis (ALS), related motor neuron disorders, asymptomatic ALS gene mutation carriers, and age-matched healthy controls[1]. Raw FASTQ files for paired-end (100bp) reverse-stranded total RNA-seq samples from iPS-derived motor neurons were obtained through the Answer ALS data portal (AALS-01184).

II. AMP-AD
   A. MSBB
      1. The Mount Sinai/JJ Peters VA Medical Center Brain Bank (MSBB – Mount Sinai NIH Neurobank) cohort consists of tissues from subjects who received a diagnosis of Alzheimer's disease or mild cognitive impairment, consistent with the Consortium to Establish a Registry for Alzheimer's disease (CERAD) protocol, as well as healthy controls[2]. From this cohort, paired-end (100bp) unstranded total RNA-seq samples from Brodmann Areas 10, 22, 36, and 44 were obtained from Synapse (syn20801188) as raw FASTQ and BAM files.
   B. ROSMAP
      1. The Religious Orders Study (ROS) and the Memory and Aging Project (MAP) are longitudinal studies of dementia and other chronic diseases of aging, where participants receive a diagnosis of Alzheimer's disease, mild cognitive impairment, or no cognitive impairment based on clinical evaluation and post mortem pathology[3]. BAM files for two batches of RNA-seq from this study were obtained from Synapse (syn23650893). The first batch consisted of paired-end (100bp) reverse-stranded polyA+ selected RNA samples sourced from the dorsolateral prefrontal cortex (DLPFC). The second batch consisted of paired-end (100bp) reverse-stranded total RNA samples sourced from the DLPFC, head of caudate nucleus, and posterior cingulate cortex.
   C. Mayo Clinic
      1. Participants in this study included subjects with Alzheimer's disease, progressive supranuclear palsy (PSP), pathological aging, and

age-matched healthy controls[4,5]. Paired-end (100bp) unstranded total RNA-seq samples from the temporal cortex (syn3163039) and cerebellum (syn5049298) were obtained from Synapse as raw FASTQ and BAM files, respectively.

III. Brain-Seq

A. The research focus of the BrainSeq Consortium is centered around neuropsychiatric disorders. Thus, from this cohort, FASTQ files for paired-end (100bp) unstranded polyA+ selected RNA-seq samples from the DLPFC of individuals with schizophrenia and healthy controls were obtained from Synapse (syn8227833)[6,7].

IV. CMC

A. The CommonMind Consortium (CMC) is a collection of postmortem brain tissue from individuals with neuropsychiatric disorders, specifically schizophrenia, bipolar disorder, and other affective disorders, as well as neurotypical controls which were received from four major collection sites[8]. Tissues from the DLPFC and anterior cingulate cortex (ACC) processed by the brain banks at Mount Sinai, University of Pennsylvania, and University of Pittsburgh collectively comprise the "CMC" cohort, from which BAM files for paired-end (100bp) total RNA-seq was obtained from Synapse (DLPFC – syn18134197; ACC – syn29442240). Additional DLPFC tissue collected at the National Institute of Mental Health Human Brain Collection Core (NIMH HBCC) make up the "HBCC" cohort, from which BAM files for paired-end (125bp) total RNA-seq samples were obtained from Synapse (syn18134197). From the "CMC" cohort, RNA-seq samples from the DLPFC were unstranded. The "HBCC" cohort and samples from the ACC were reverse-stranded.

V. GTEx

A. The Adult Genotype Tissue Expression (GTEx v8) project collects samples from non-diseased tissues[9]. We obtained paired-end (75bp) unstranded polyA+ selected RNA-seq samples which were sourced from 13 brain regions from the database of Genotypes and Phenotypes (dbGaP)(phs000424.v8).

VI. Knight ADRC

A. Paired-end (125bp) total RNA-seq samples from the DLPFC of Alzheimer's disease patients and controls which were collected by the Knight Alzheimer's

Disease Research Center (ADRC) were downloaded from NIAGADS (NG00083)[10].

VII. NABEC

A. We obtained paired-end (100bp) unstranded total RNA-seq samples from neurotypical brain donors as raw FASTQ files from the North American Brain Expression Consortium (NABEC) through dbGaP (phs001300.v2.p1).

VIII. NYGC ALS

A. The New York Genome Center (NYGC) ALS consortium is a data-sharing effort which brings together 45 institutions collecting central nervous system (CNS) as well as peripheral tissue samples from subjects with a spectrum of ALS and related movement disorders, as well as other neuropathologies including neurodegenerative diseases such as Alzheimer's disease and frontotemporal dementia[11]. As consortium members, we were granted access to raw FASTQ files from several batches of paired-end (100bp and 125bp) total RNA-seq from 11 CNS tissues published (GSE137810) and unpublished, contributed from 9 medical centers.

IX. UKBEC

A. We obtained raw FASTQ files of paired-end (100bp) unstranded total RNA-seq samples from the substantia nigra and putamen of neurotypical brain donors originating from the United Kingdom Brain Expression Consortium (UKBEC) dataset from the European Genome-Phenome Archive (EGAS00001002113).

For all primary research studies listed above, the permission to collect human brain material was obtained from the Ethical Committee of Institutional Review Boards (IRB). Informed consent for autopsy, the use of the brain tissue, and accompanied clinical information for research purposes was obtained per donor ante-mortem. All autopsies were performed with written consent from the legal next-of-kin. The study was performed under IRB-approved guidance and regulations to keep all patient information strictly de-identified. All research conformed to the principles of the Helsinki Declaration. Data were downloaded from the Synapse AD Knowledge Portal, PsychENCODE Portal, CommonMind Consortium Knowledge Portal, BrainSeq Consortium, NYGC ALS Consortium, KnightADRC (shared by Dr. Carlos Cruchaga), and GTEx (from dbGaP). All protected data were accessed via dbGaP. We thank the study participants of all the BigBrain cohorts for their generous gifts of brain donation, without which none of this research would be possible.

### Sample sizes in BigBrain QTL studies

Meta-analyses of RNA editing (edQTL), gene expression (eQTL), and RNA splicing (sQTL) were all performed as part of the BigBrain Project. Sample size discrepancies between *Réal et al.* (n=10,725 samples, n=4,656 individuals) and *Dredge et al.* (n=10,202 samples, n=4,471 individuals) are due to criteria necessary for calling high-confidence RNA editing.

### Pre-processing RNA-sequencing data

RNA-seq samples obtained from various studies came in two different formats: raw FASTQ files or mapped BAM files (see above and **Supplementary Table 1**). In the case of raw FASTQ files, no additional pre-processing took place. In the case of mapped BAM files, the data was reverted to FASTQ format using either Bazam[12] (unstranded data) or Picard v.3.1.1 SamToFastq (stranded data).

### Mapping RNA-sequencing data

Raw and converted FASTQ files were uniformly processed using the RAPiD pipeline, whereby STAR[13] v.2.7.2a mapped RNA-seq samples to the hg38 genome build with GENCODE v.30 transcriptome reference. Transcript expression was measured by RSEM[14] v.1.3.1 and summarized to a gene level with tximport[15] v.1.18. Picard v.2.20.3 was leveraged to determine per sample quality control metrics. Genes with at least 1 count per million in at least 10% of samples were kept and VOOM normalized by limma[16].

### Genetic quality control

Genetic data came in the form of whole-genome sequencing (WGS) or SNP arrays (**Supplementary Table 2**). Quality control (QC) used an in-house pipeline built around PLINK[17].

In the case of SNP arrays, bcftools v.1.9 and VCFtools v.0.1.15 were used to remove variants with call rates < 95%, minor allele frequency (MAF) < 5%, and Hardy-Weinberg equilibrium P < 1e-6. The call rate for each sample had to be > 95% for it to be retained. Genotypes were then imputed with the Michigan Imputation Server[18] v.1.4.1 with the 1000 Genomes Project[19] (Phase 3) v.5 (GRCh37) European panel and phased with Eagle v.2.4 in quality control and imputation mode with the “rsq” filter set to 0.3. Picard’s liftoverVCF and the “b37ToHg38.over.chain.gz” liftover chain file were used to lift over variants from GRCh37 to GRCh38 post-imputation, in order to match the RNA-seq samples. A final round of QC was applied to the post-imputation variants to remove indels and multiallelic SNPs and only retain SNPs with MAF > 5% and Hardy-Weinberg equilibrium P > 1e-6.

In the case of WGS, a Variant Quality Score Recalibration (VQSR) model was trained using GATK's VariantRecalibrator and this model was then applied to the joint call-set with ApplyRecalibration truth sensitivity levels of 99.8% for SNPs and 99% for indels. Variants that failed VSQR and Low Complexity Region (LCR) were excluded during WGS QC. The following filters were applied to the final set of variants: not overlapping hg38 blacklisted regions; genotype level read depth (GDP) > 10; genotype quality (GQ) > 20; missingness < 15%; total read depth > 5000 (~10x coverage); 58.75 > mapping quality (MQ) > 61.25; variant quality score log-odds (VQSLOD) > 7.81; and inbreeding coefficient > -0.8. Sample-level QC removed samples with a mean depth < 0.2 and call rate < 0.99. Further, samples were removed if the inferred sex was discordant with that recorded in the study metadata.

Finally, all VCF files were filtered to exclude singletons and resolve incongruent SNP IDs across cohorts. The final number of variants retained in the VCF files varied between cohorts, reflecting cohort-specific genetic architectures and filtering stringency (**Supplementary Table 2**). RNA-seq samples were matched to the donor of origin using the tool MBV from QTLtools[20] v.1.3 and genetic ancestry was inferred by Somalier[21]. Samples which had a duplicated or up to third-degree-relation or pairwise kinship coefficient estimated using KING[22] v.2.2.5 were removed.

### QTL mapping and meta-analysis with mmQTL

Normalized gene expression, splice junction usage, or RNA editing ratio matrices were scaled and centered to a mean of 0 and standard deviation of 1, and quantile normalized. PEER factors were estimated from the phenotypes to identify hidden covariates. Age was used where metadata was available for individual studies (excluding Answer ALS and NABEC). Biological sex was inferred by Somalier[21] based on scaled mean depth of sites on chrX and chrY.

Using mmQTL[23] v.26a, a linear mixed model was applied for QTL mapping and all 43 datasets were combined with a random-effects meta-analysis. Briefly, genotypes were first converted into Plink v.2.3 format and then a genotype relatedness matrix was constructed using GCTA[24] v.1.94.1. With a MAF cutoff of 0.01, all SNPs within a 1 Mb window were tested for association with respective phenotypes. The resultant meta-analysis random effects P-values are first Bonferroni adjusted for the number of SNPs tested per feature, then the Benjamini-Hochberg method is applied to control false discovery rate against the total number of features tested.

### Fine-mapping

PolyFun[26] was used for functionally informed fine-mapping, with SuSiE as the fine-mapping engine[26–28]. Top QTLs were selected from full mmQTL summary statistics. SNP-level summary statistics were processed with PolyFun's munge_polyfun_sumstats.py, modified to drop variants that were zero or missing across all tissue-cohort pairs. To mitigate variance differences across tissues, fixed Z-scores were replaced with random Z-scores whose signs matched the original fixed Z's.

In-sample LD was computed from 3,884 European-ancestry participants in the BigBrain study to match the genotypes underlying the QTLs statistics. Of 9,130,833 tested SNPs, variants observed in ≥21 tissue-cohort pairs (51% of pairs) were retained, yielding 7,375,120 widely shared SNPs (81%) for LD. VCFs from multiple cohorts were converted to PLINK format (.bed/.bim/.fam) using PLINK v1.9, and LD matrices were computed per chromosome for efficiency.

Fine-mapping was run with PolyFun's finemapper.py in 1 Mb windows around both the start and end of each junction (for sQTLs), start of the gene (for eQTLs) and the editing site (for edQTLs). SuSiE was configured with L=5 (maximum causal variants per locus) and the following options: --no-sort-pip, --allow-swapped-indel-alleles, and --susie-resvar 1. Analyses were executed per chromosome and merged downstream.

### Mediated expression score regression

QTL sets were applied to explain disease heritability from Alzheimer's disease[29], amyotrophic lateral sclerosis[30], bipolar disorder[31], major depressive disorder[32], multiple sclerosis[33], Parkinson's disease[34], and schizophrenia[35] using mediated expression score regression (MESC[36]). MESC estimates the proportion of common variant disease heritability ($h^2_g$) that is mediated through a set of QTLs ($h^2_{med}$). The most recent height GWAS[37] was included as a control. In the case of Alzheimer's disease, the APOE locus was excluded.

### Colocalization analysis

COLOC[38] was used to test whether SNPs from Alzheimer's disease[29,39–42], ALS[30,43], bipolar disorder[31,44], Parkinson's disease[34], or schizophrenia[35,45] are colocalized with QTLs. For each GWAS, the nominal summary statistics were extracted for SNPs within a 1 Mb window of the index SNP at all genome-wide significant loci, excluding loci overlapping the human major histocompatibility complex (hg19 chr6:28,477,797-33,448,354) and the microtubule-associated protein tau H1/H2 haplotype region (hg19 chr17:43,628,944-44,571,603). Colocalizations were only considered for genome-wide significant SNPs from GWAS (p-value < $5e^{-8}$) and where the

GWAS and QTL lead SNPs were within 100 Kb of each other or within 500 Kb and in moderate LD ($r^2 > 0.1$). We define strong evidence of colocalization at posterior probability of hypothesis 4 (PP4) > 0.8. The 1000 Genomes (Phase 3) European populations were used by LDLinkR v.1.1.2 to determine linkage disequilibrium between GWAS and QTL SNPs. In the case where multiple GWAS of the same disease were used, a consensus set of loci was determined, thus colocalizations were not double-counted.

### References

1. Baxi, E. G. *et al.* Answer ALS, a large-scale resource for sporadic and familial ALS combining clinical and multi-omics data from induced pluripotent cell lines. *Nat. Neurosci.* **25**, 226–+ (2022).
2. Wang, M. H. *et al.* The Mount Sinai cohort of large-scale genomic, transcriptomic and proteomic data in Alzheimer's disease. *Sci. Data* **5**, (2018).
3. Bennett, D. A. *et al.* Selected Findings from the Religious Orders Study and Rush Memory and Aging Project. *JOURNAL OF ALZHEIMERS DISEASE* **33**, S397–S403 (2013).
4. Allen, M. *et al.* Human whole genome genotype and transcriptome data for Alzheimer's and other neurodegenerative diseases. *Sci. Data* **3**, (2016).
5. Carrasquillo, M. M. *et al.* Genetic variation in *PCDH11X* is associated with susceptibility to late-onset Alzheimer's disease. *Nat. Genet.* **41**, 192–198 (2009).
6. Schubert, C. R. *et al.* BrainSeq: Neurogenomics to Drive Novel Target Discovery for Neuropsychiatric Disorders. *Neuron* **88**, 1078–1083 (2015).
7. Jaffe, A. E. *et al.* Developmental and genetic regulation of the human cortex transcriptome illuminate schizophrenia pathogenesis. *Nat. Neurosci.* **21**, 1117–+ (2018).
8. Hoffman, G. E. *et al.* CommonMind Consortium provides transcriptomic and epigenomic data for Schizophrenia and Bipolar Disorder. *Sci. Data* **6**, (2019).
9. Aguet, F. *et al.* The GTEx Consortium atlas of genetic regulatory effects across human tissues. *SCIENCE* **369**, 1318–1330 (2020).
10. Dube, U. *et al.* An atlas of cortical circular RNA expression in Alzheimer disease brains demonstrates clinical and pathological associations. *Nat. Neurosci.* **22**, 1903–+ (2019).
11. Humphrey, J. *et al.* Integrative transcriptomic analysis of the amyotrophic lateral sclerosis spinal cord implicates glial activation and suggests new risk genes. *Nat. Neurosci.* **26**, 150–+ (2023).

12. Sadedin, S. P. & Oshlack, A. Bazam: a rapid method for read extraction and realignment of high-throughput sequencing data. *Genome Biol.* **20**, (2019).
13. Dobin, A. *et al.* STAR: ultrafast universal RNA-seq aligner. *Bioinformatics* **29**, 15–21 (2013).
14. Li, B. & Dewey, C. N. RSEM: accurate transcript quantification from RNA-Seq data with or without a reference genome. *BMC Bioinformatics* **12**, (2011).
15. Soneson, C., Love, M. I. & Robinson, M. D. Differential analyses for RNA-seq: transcript-level estimates improve gene-level inferences. *F1000Res.* **4**, 1521 (2015).
16. Ritchie, M. E. *et al.* *limma* powers differential expression analyses for RNA-sequencing and microarray studies. *Nucleic Acids Res.* **43**, (2015).
17. Purcell, S. *et al.* PLINK: A tool set for whole-genome association and population-based linkage analyses. *Am. J. Hum. Genet.* **81**, 559–575 (2007).
18. Das, S. *et al.* Next-generation genotype imputation service and methods. *Nat. Genet.* **48**, 1284–1287 (2016).
19. Altshuler, D. M. *et al.* A global reference for human genetic variation. *NATURE* **526**, 68–+ (2015).
20. Fort, A. *et al.* *MBV*: a method to solve sample mislabeling and detect technical bias in large combined genotype and sequencing assay datasets. *Bioinformatics* **33**, 1895–1897 (2017).
21. Pedersen, B. S. *et al.* Somalier: rapid relatedness estimation for cancer and germline studies using efficient genome sketches. *Genome Med.* **12**, (2020).
22. Manichaikul, A. *et al.* Robust relationship inference in genome-wide association studies. *Bioinformatics* **26**, 2867–2873 (2010).
23. Zeng, B. A. *et al.* Multi-ancestry eQTL meta-analysis of human brain identifies candidate causal variants for brain-related traits. *Nat. Genet.* **54**, 161–+ (2022).
24. Yang, J. A., Lee, S. H., Goddard, M. E. & Visscher, P. M. GCTA: A Tool for Genome-wide Complex Trait Analysis. *Am. J. Hum. Genet.* **88**, 76–82 (2011).
25. Storey, J. D. The positive false discovery rate:: A Bayesian interpretation and the *q*-value.

*Ann. Stat.* **31**, 2013–2035 (2003).

26. Weissbrod, O. *et al.* Functionally informed fine-mapping and polygenic localization of complex trait heritability. *Nat. Genet.* **52**, 1355–1363 (2020).
27. Wang, G., Sarkar, A., Carbonetto, P. & Stephens, M. A simple new approach to variable selection in regression, with application to genetic fine mapping. *J. R. Stat. Soc. Series B Stat. Methodol.* **82**, 1273–1300 (2020).
28. Zou, Y., Carbonetto, P., Wang, G. & Stephens, M. Fine-mapping from summary data with the 'Sum of Single Effects' model. *PLoS Genet.* **18**, e1010299 (2022).
29. Bellenguez, C. *et al.* New insights into the genetic etiology of Alzheimer's disease and related dementias. *Nat. Genet.* **54**, 412–436 (2022).
30. van Rheenen, W. *et al.* Author Correction: Common and rare variant association analyses in amyotrophic lateral sclerosis identify 15 risk loci with distinct genetic architectures and neuron-specific biology. *Nat. Genet.* **54**, 361 (2022).
31. Stahl, E. A. *et al.* Genome-wide association study identifies 30 loci associated with bipolar disorder. *Nat. Genet.* **51**, 793–803 (2019).
32. Wray, N. R. *et al.* Genome-wide association analyses identify 44 risk variants and refine the genetic architecture of major depression. *Nat. Genet.* **50**, 668–681 (2018).
33. International Multiple Sclerosis Genetics Consortium. Multiple sclerosis genomic map implicates peripheral immune cells and microglia in susceptibility. *Science* **365**, eaav7188 (2019).
34. Nalls, M. A. *et al.* Identification of novel risk loci, causal insights, and heritable risk for Parkinson's disease: a meta-analysis of genome-wide association studies. *Lancet Neurol.* **18**, 1091–1102 (2019).
35. Schizophrenia Working Group of the Psychiatric Genomics Consortium. Biological insights from 108 schizophrenia-associated genetic loci. *Nature* **511**, 421–427 (2014).
36. Yao, D. W., O'Connor, L. J., Price, A. L. & Gusev, A. Quantifying genetic effects on disease

mediated by assayed gene expression levels. *Nat. Genet.* **52**, 626–633 (2020).

37. Yengo, L. *et al.* A saturated map of common genetic variants associated with human height. *Nature* **610**, 704–712 (2022).

38. Wallace, C. Statistical Testing of Shared Genetic Control for Potentially Related Traits. *Genet. Epidemiol.* **37**, 802–813 (2013).

39. Jansen, I. E. *et al.* Author Correction: Genome-wide meta-analysis identifies new loci and functional pathways influencing Alzheimer's disease risk. *Nat. Genet.* **52**, 354 (2020).

40. Kunkle, B. W. *et al.* Genetic meta-analysis of diagnosed Alzheimer's disease identifies new risk loci and implicates Aβ, tau, immunity and lipid processing. *Nat. Genet.* **51**, 414–430 (2019).

41. Marioni, R. E. *et al.* Correction: GWAS on family history of Alzheimer's disease. *Transl. Psychiatry* **9**, 161 (2019).

42. Lambert, J. C. *et al.* Meta-analysis of 74,046 individuals identifies 11 new susceptibility loci for Alzheimer's disease. *Nat. Genet.* **45**, 1452–1458 (2013).

43. Nicolas, A. *et al.* Genome-wide analyses identify KIF5A as a novel ALS gene. *Neuron* **97**, 1268–1283.e6 (2018).

44. Mullins, N. *et al.* Genome-wide association study of more than 40,000 bipolar disorder cases provides new insights into the underlying biology. *Nat. Genet.* **53**, 817–829 (2021).

45. Trubetskoy, V. *et al.* Mapping genomic loci implicates genes and synaptic biology in schizophrenia. *Nature* **604**, 502–508 (2022).
