## Supplementary figures for "Mapping genetic effects on splicing in ten thousand post-mortem brain samples reveals novel mediators of neurological disease risk"

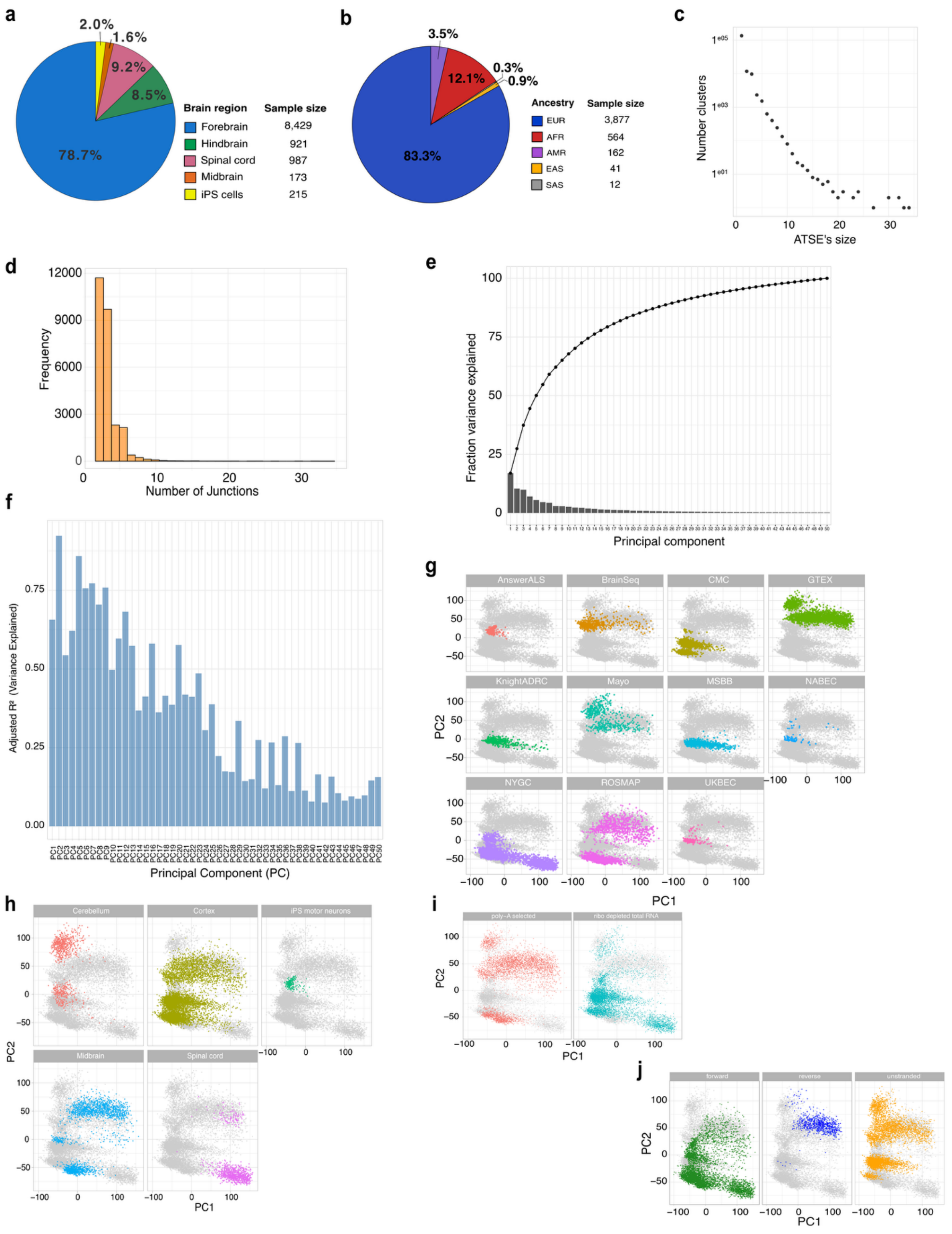


**S1: (a)** Pie chart showing the distribution of RNA-seq samples by brain region and iPSC cell type. Each colored wedge represents a tissue category, annotated with percentage values and matched to a legend indicating sample counts. **(b)** Pie chart displaying ancestry proportions of RNA-seq samples. Segments are color-coded by ancestry group, with corresponding percentage labels and a legend listing sample sizes per group. **(c)** Scatter plot displaying the number of clusters (y-axis, log-scaled) as a function of ATSE size (x-axis). Each point represents a unique ATSE cluster size and its corresponding count. **(d)** Histogram showing the distribution of junction counts per ATSE. The x-axis indicates the number of junctions per cluster, and the y-axis shows frequency. Bars are shaded in orange. **(e)** Bar plot showing the proportion of variance explained by each principal component from PCA. The x-axis shows principal component number; the y-axis indicates fraction of variance explained. A cumulative variance line with black dots connects the bars to visualize total variance explained. **(f)** Bar plot showing adjusted $R^2$ values (y-axis) for the first 50 principal components (x-axis), representing the proportion of variance in each PC explained jointly by brain region and cohort. **(g)** Grid of 12 scatter plots showing principal component 1 (PC1) versus PC2 projections. Each panel corresponds to a different cohort, with samples colored by cohort identity and background samples in gray. **(h)** Grid of 6 scatter plots showing PC1 vs. PC2 colored by brain region. Each subplot highlights one region with colored points, with remaining samples in gray. **(i)** Two-panel scatter plot of PC1 vs. PC2 colored by RNA library preparation type (polyA-selected vs. ribo-depleted), with gray background points. **(j)** Three-panel scatter plot of PC1 vs. PC2 colored by strand orientation category (forward, reverse, or unstranded), with gray background samples.

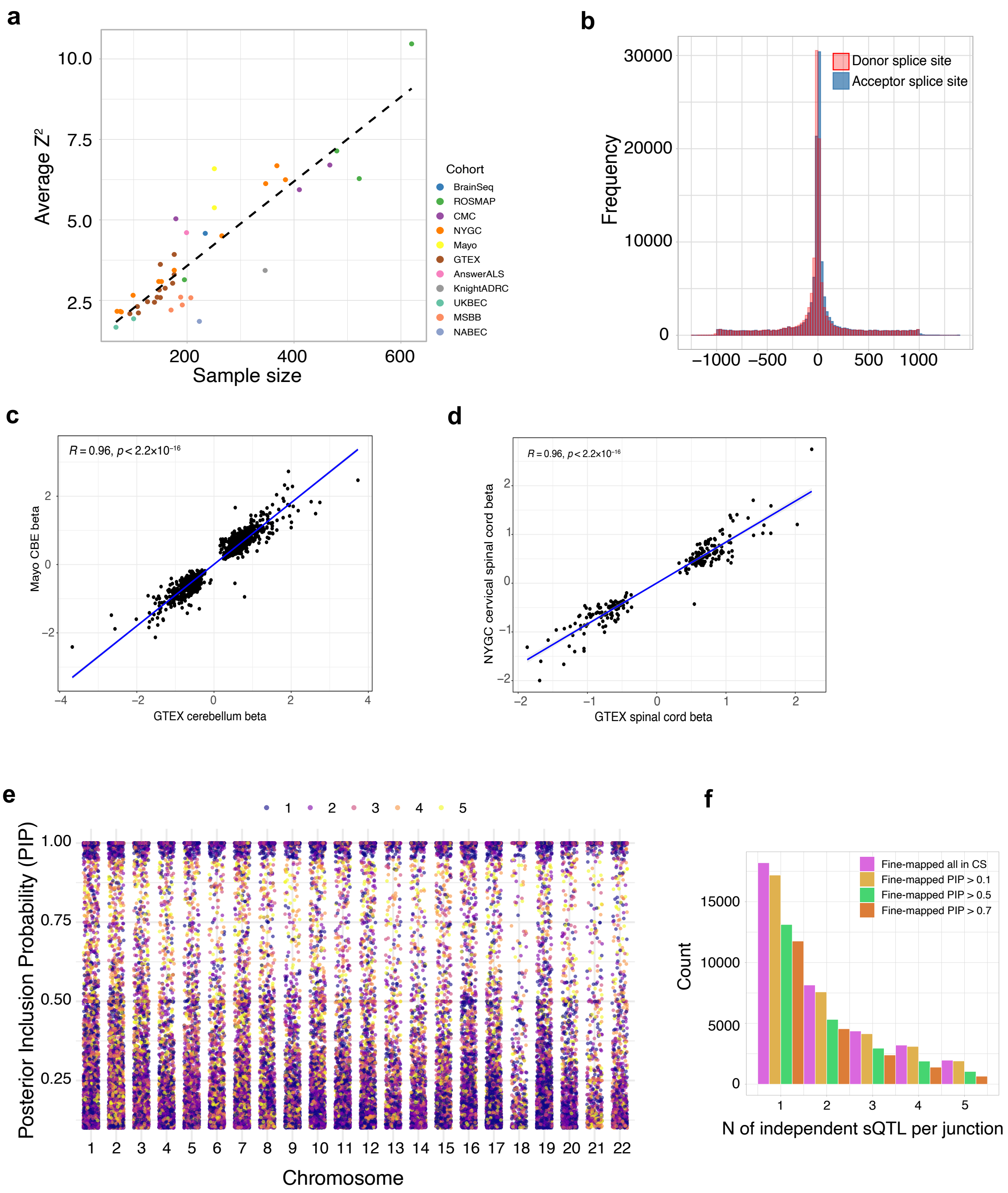


**S2: (a)** Scatter plot of average Z-square scores versus sample size for each cohort, with points colored by cohort. A dashed black line represents the expected relationship under null variance scaling. **(b)** Histogram of sQTL-SNP distances from donor (red) and acceptor (blue) splice sites across all tested variants. **(c,d)** Scatter plots comparing meta-analysis Z-scores to individual cohort Z-scores for (d) Mayo CBE and GTEx cerebellum; (d) NYGC cervical spinal cord and GTEX spinal cord . Each point represents a tested junction-SNP pair, with a blue regression line and

$R^2$/p-value indicating agreement. **(e)** Manhattan-like strip plot showing posterior inclusion probabilities (PIPs) from SuSiE fine-mapping across autosomal chromosomes (x-axis). Each point represents a variant, positioned by chromosome and colored by 95% credible set membership. Points are jittered horizontally to aid visualization of PIP distributions (y-axis). **(f)** Number of independent fine-mapped sQTLs per junction, stratified by posterior inclusion probability (PIP) thresholds.

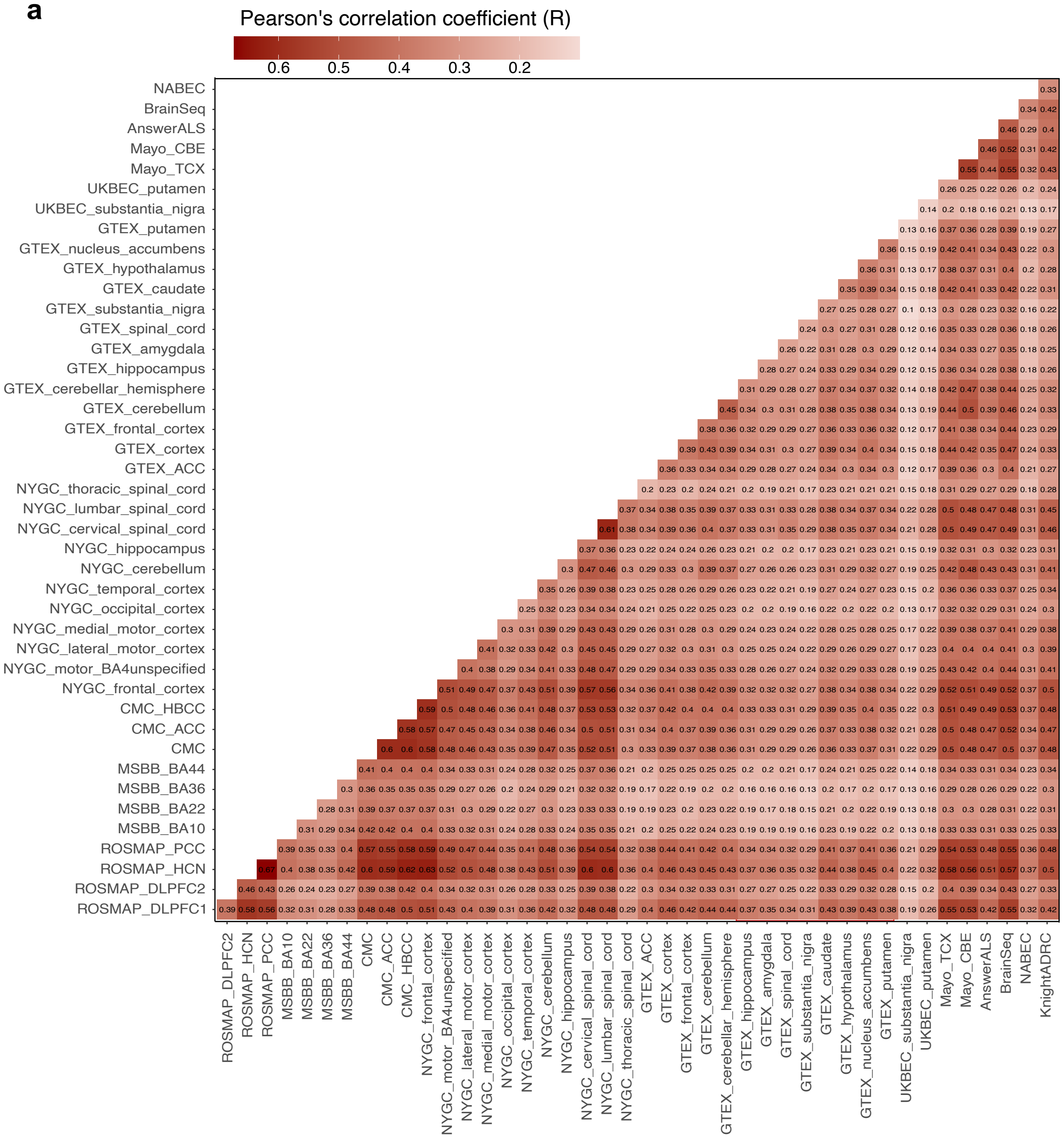


**S3: (a)** Heatmap of pairwise Pearson correlation coefficients (R) of junction-level sQTL Z-scores between tissue-cohort pairs.

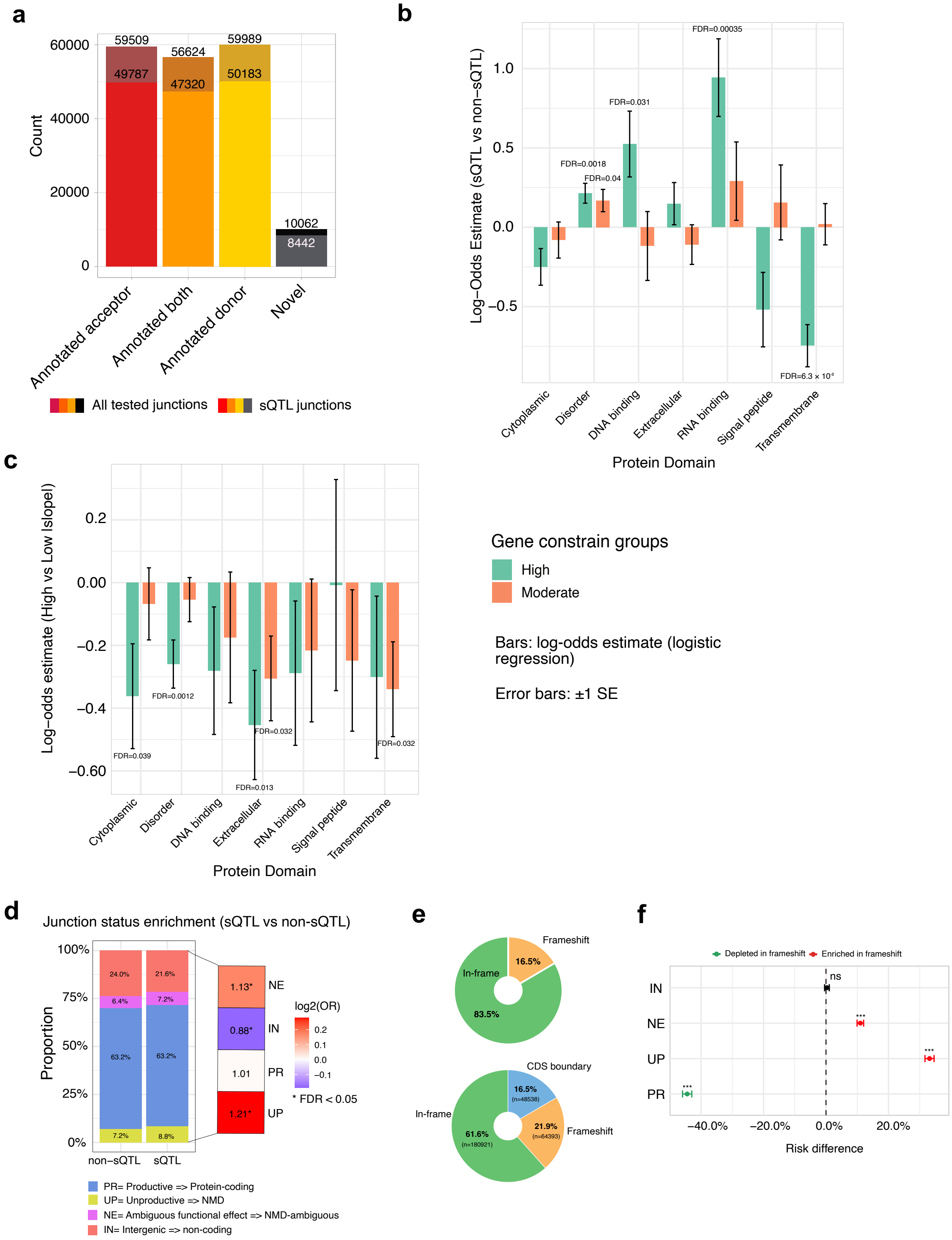


**S4: (a)** Bar plot showing the number of tested junctions (light bars) and significant sQTL junctions (dark bars) stratified by annotation category: junctions with annotated acceptor, donor, both splice sites, or novel junctions. **(b)** Logistic regression modeling the likelihood that a junction is an sQTL (vs. non-sQTL), based on protein domain overlap and gene constraint (GeneBayes). Bars show log-odds estimates (±1 SE) from a logistic model predicting sQTL vs non-sQTL as a function of protein-domain overlap, stratified by GeneBayes constraint (High = green, Moderate = orange). Positive bars indicate enrichment of sQTLs among junctions overlapping that domain; negative bars indicate depletion. Text above bars reports the BH-FDR for the coefficient (omitted when not significant). The zero line denotes no effect. **(c)** Bars show log-odds estimates (±1 SE) from a logistic model predicting high-effect sQTLs (top-decile $|\beta|$; non-sQTLs set to $\beta=0$) vs low as a function of protein-domain overlap, again stratified by GeneBayes constraint (High = green, Moderate = orange). Positive bars indicate enrichment among high-effect events; negative bars indicate depletion. Text labels give BH-FDR for the domain effect; the zero line marks no effect. **(d)** Stacked bar plot showing the distribution of predicted junction consequences (productive, unproductive, NMD, non-coding, ambiguous) between sQTLs and non-sQTLs. Log odds ratios (right) highlight significant enrichment of unproductive splicing (UP) and depletion in ambiguous (NE) categories among sQTLs (FDR < 0.05 indicated by *). **(e)** Donut charts summarizing frame consequences for CDS-overlapping sQTLs. Top: Per-junction (collapsed) labels after aggregating over all annotated transcripts; a junction is called in-frame or frameshift only when non-boundary evidence agrees across transcripts. Bottom: Per-transcript calls (counts of junction×transcript pairs before collapsing). The additional “CDS boundary” wedge marks cases with zero CDS overlap but adjacency to a CDS edge (boundary-only; see Methods). **(f)** Frameshift vs in-frame difference for each class (PR, UP, NE, IN). Points show estimates with 95% CIs from a two-sample proportion test ($\chi^2$ with Yates). Red = enriched, green = depleted; asterisks denote BH-FDR < 0.05.

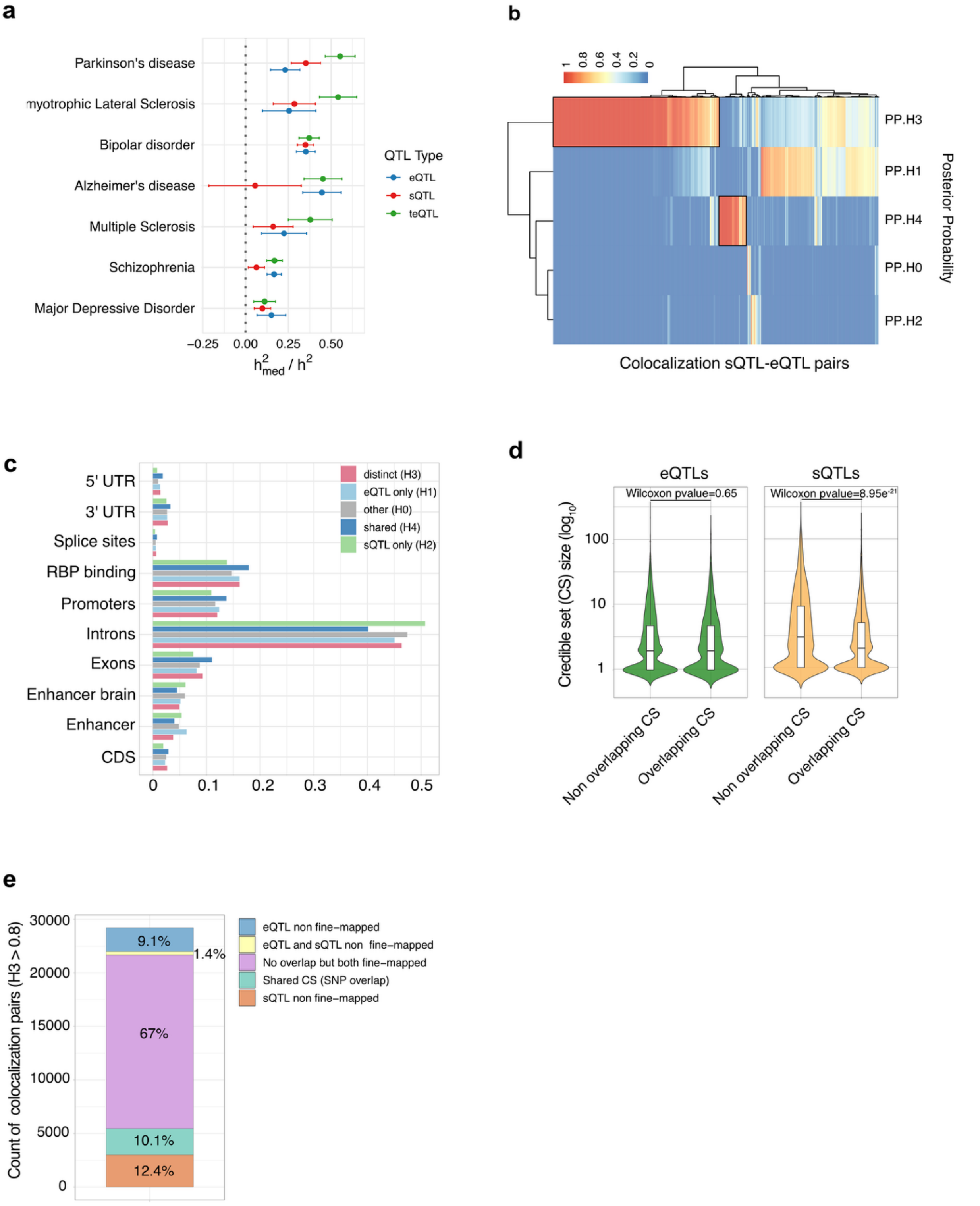


**S5: (a)** Proportion of GWAS heritability explained by fine-mapped QTLs (eQTLs in blue, sQTLs in red and transcript QTL (teQTL) in green) for six neurological GWAS traits. **(b)** Heatmap showing posterior probabilities (PP.H0-H4) for colocalization between sQTL and eQTL signals across all tested loci, with hierarchical clustering. **(c)** Barplot showing the proportion of colocalization outcomes across genomic annotations. Each annotation (e.g., promoters, splice sites, RBP binding regions, introns, exons, etc.) is stratified by colocalization posterior probability categories: shared causal variant (H4), sQTL-specific signal (H2), eQTL-specific signal (H1), distinct causal variants (H3), and no association (H0). **(d)** Violin plot comparing the size of fine-mapped credible sets (CS) between colocalized and non-colocalized signals, separately for eQTLs (green) and sQTLs (light orange). CS sizes are defined as the number of variants with posterior inclusion probability (PIP) ≥ 0.1. Colocalized signals have smaller CS sizes, indicating more precise fine-mapping. **(e)** Barplot classifying loci with distinct regulatory signals (PP.H3 ≥ 0.8) from sQTL-eQTL colocalization analysis. Each bar shows the proportion of loci where the fine-mapped credible sets (PIP ≥ 0.1) are: (i) shared between sQTL and eQTL (SNP overlap), (ii) partially fine-mapped (credible set present in only one QTL type), or (iii) not fine-mapped in either. This breakdown reveals the extent to which distinct signals identified by colocalization are supported by variant-level resolution from fine-mapping.

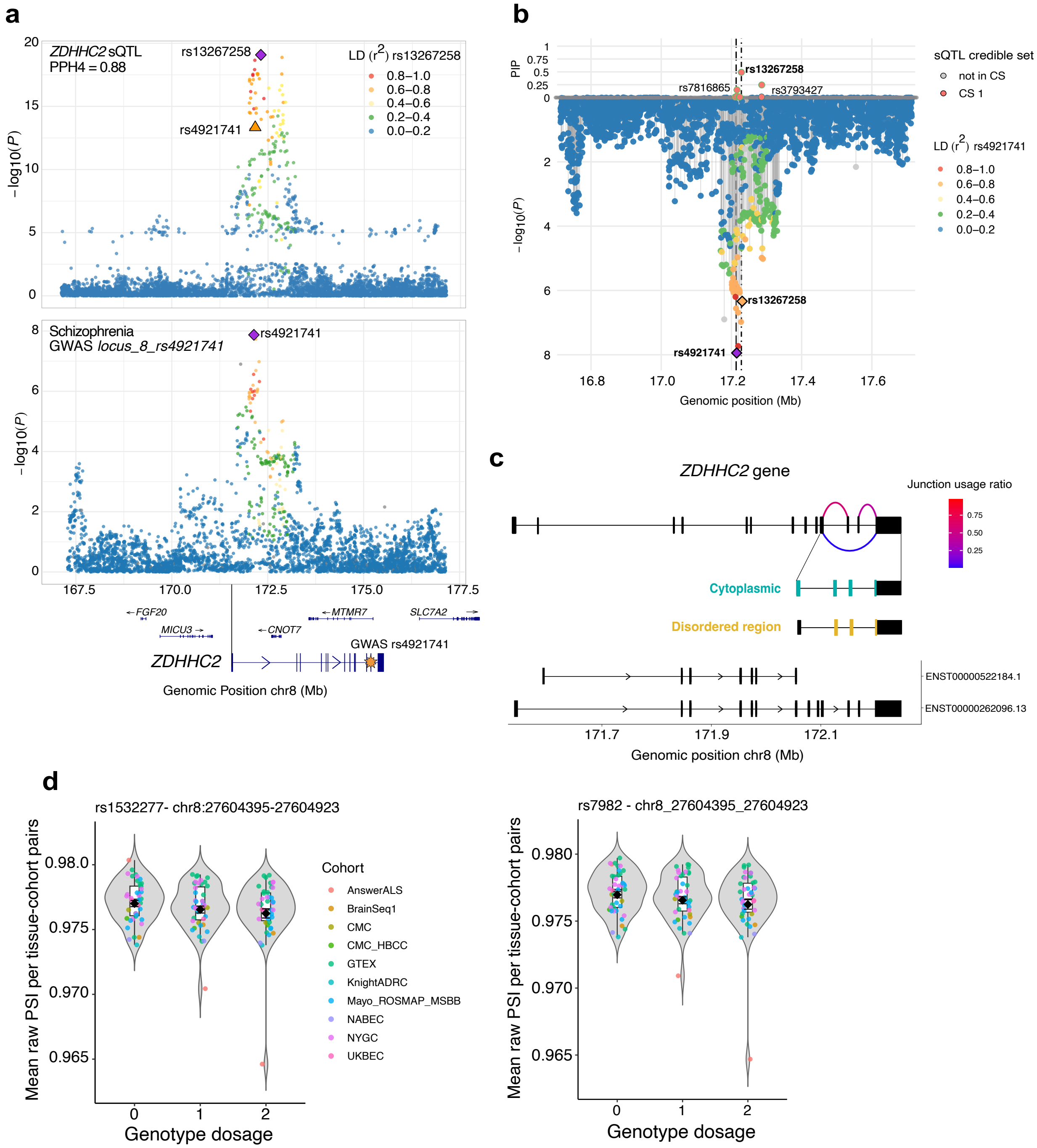


**S6: (a)** Colocalization of *ZDHHC2* sQTL and schizophrenia GWAS signals. Top: sQTL $-\log_{10}(P)$ values for *ZDHHC2* junction usage, with lead fine-mapped variant rs13267258 (purple diamond) and LD with rs13267258 shown by color. Bottom: GWAS $-\log_{10}(P)$ values at the schizophrenia locus near rs49217141 (orange triangle), highlighting the overlapping signal. **(b)** Mirror plot summarizing sQTL fine-mapping and association. Top: SuSiE PIP. Bottom: sQTL $-\log_{10}(P)$. Each point is a SNP; fill encodes LD ($r^2$) to the GWAS index rs4921741. Variants in the 95% credible set are highlighted as CS1 (red); open circles are not in the CS. Diamonds mark index variants (orange = sQTL rs13267258, purple = GWAS rs4921741). Dashed vertical lines indicate their genomic positions. **(c)** Gene structure and domain annotation for *ZDHHC2*. Top: arcs indicate splice junctions colored by junction usage ratio (PSI). Overlapping UniProt-annotated domains are shown below: cytoplasmic (blue) and disordered regions (yellow). Bottom: Ensembl transcript models for *ZDHHC2* (hg38). **(d)** Violin plot for the genotype-PSI relationships for rs1532277 and rs7982 across cohorts and tissues, shown separately for the two tandem-acceptor junctions. PSI shown are raw PSI before scale-center, quantile normalization and PEERs correction. Each violin summarizes the distribution of per tissue-cohort mean PSI at each genotype dosage (0/1/2); boxes show median and IQR; points are individual tissue-cohort means; black diamonds mark the sample-size-weighted mean (± SE). Left (chr8:27604395–27604923-rs1532277, ΔE): PSI increases with risk-allele dosage ($\beta > 0$). Right (chr8:27604392–27604923-rs7982, E-retained): PSI decreases with risk-allele dosage ($\beta < 0$).
